# A Habenula Neural Biomarker Simultaneously Tracks Weekly and Daily Symptom Variations during Deep Brain Stimulation Therapy for Depression

**DOI:** 10.1101/2024.07.29.24310966

**Authors:** Shi Liu, Yu Qi, Shaohua Hu, Ning Wei, Jianmin Zhang, Junming Zhu, Hemmings Wu, Hailan Hu, Yuxiao Yang, Yueming Wang

## Abstract

Deep brain stimulation (DBS) targeting the lateral habenula (LHb) is a promising therapy for treatment-resistant depression (TRD) but its clinical effect has been variable, which can be improved by adaptive DBS (aDBS) guided by a neural biomarker of depression symptoms. A clinically-viable neural biomarker is desired to classify depression symptom states, track both slow and fast symptom variations during the treatment, respond to DBS parameter alterations, and be neurobiologically interpretable, which is currently lacking. Here, we conducted a study on one TRD patient who achieved remission following a 41-week LHb DBS treatment, during which we assessed slow symptom variations using weekly clinical ratings and fast variations using daily self-reports. We recorded daily LHb local field potentials (LFP) concurrently with the reports during the entire treatment process. We then used machine learning methods to identify a personalized depression neural biomarker from spectral and temporal LFP features. The identified neural biomarker classified high and low depression symptom severity states with a cross-validated accuracy of 0.97, with the most contributing spectral and temporal feature being LFP beta band power and Hurst exponent, respectively. It further simultaneously tracked both weekly (slow) and daily (fast) depression symptom variation dynamics, achieving test data explained variance of 0.74 and 0.63, respectively. It also responded to DBS frequency alterations. Finally, it can be neurobiologically interpreted as indicating changes in LHb excitatory and inhibitory balance during DBS treatment. Together, our results hold promise to identify clinically-viable neural biomarkers to facilitate future aDBS for treating TRD.

## 1. Introduction

Major depressive disorder (MDD) is one of the most common neuropsychiatric disorders, affecting over 300 million individuals worldwide [1] . Approximately 30^7^ of MDD patients are treatment-resistant, meaning they do not respond adequately to at least two antidepressant trials [2]. Deep brain stimulation (DBS) is a neurosurgical procedure that allows targeted circuit-based neuromodulation [3]. It has emerged as a promising treatment option for patients with treatment-resistant depression (TRD) [4–6], as shown by open-label studies targeting various brain structures involved in the brain’s “reward” system that mediates positive motivations. Such targets include the subcallosal cingulate cortex (SCC) [7], the ventral capsule/ventral striatum (VC/VS) [8], the medial forebrain bundle (MFB) [9], and the bed nucleus of the stria terminalis (BNST) [10]. However, several recent double-blinded clinical trials have shown that the effects of DBS targeting these brain structures are inconsistent across patients [11–15]. As a potential improvement over DBS, adaptive DBS (aDBS) optimizes DBS parameters in real-time by using neural signals as feedback for enhancing clinical efficacy [16]. A recent study implements aDBS targeting VC/VS in a TRD patient by triggering stimulation only when the local field potential (LFP) signal pattern indicates worsening of depression symptoms, achieving rapid alleviation of depression symptoms [17].

The lateral habenula (LHb) is a hub structure that plays a central role in the brain’s “anti-reward” system that mediates negative motivations [18–20]. Animal studies have systematically shown that the local bursting firing patterns in LHb are closely related to depression-like behaviors and that neuromodulation of LHb has significant antidepressant effects [21–23]. Several clinical studies have reported single-patient depression symptom alleviation following LHb DBS since 2010 [24–27]. On the other hand, two recent clinical studies on seven or six patients has shown more variable effects of LHb DBS across patients [28,29]. Similar to other DBS targets, aDBS for LHb also provides a promising path towards improved and more consistent treatment effects across TRD patients.

A critical and fundamental requirement for developing LHb aDBS is the identification of an LHb neural biomarker of depression symptoms during the DBS treatment to provide the necessary feedback signal [30,31]. A population-level SCC LFP spectral power biomarker has been identified for tracking depression symptom recovery with SCC DBS in five TRD patients [32]. Personalized amygdala and BNST LFP gamma power biomarkers have been identified for optimizing VC/VS DBS [33]. For LHb DBS, LFP signals have been recorded before the DBS treatment starts but not during the multi-month-long treatment process [28,29,34] and several studies have found statistical correlations between pre-treatment LHb LFP spectral features and after-treatment depression symptom ratings [28,29,34]. However, it is unknown whether the identified LFP features can classify depression symptom severity states or track the temporal dynamics of depression symptom variations during the DBS treatment process. Therefore, a useful neural biomarker for realizing LHb aDBS is still lacking.

A clinically-viable neural biomarker is desired to be able to track both the slow and fast temporal dynamics of depression symptom variations during DBS. This is because both natural and DBS-induced depression symptom changes can vary at different time scales, with both slow-changing dynamics over months or weeks [35–39] and fast-changing dynamics over hours or days [9,40–43]. Existing neural biomarker studies have focused on tracking the temporal dynamics of either slow or fast symptom variations. The aforementioned SCC neural biomarker for SCC DBS tracks the temporal dynamics of the weekly symptom variations over 24 weeks [32]. The aforementioned amygdala and BNST neural biomarkers for VC/VS DBS track the faster temporal dynamics of symptom variations within several days [17,33]. Several other studies have also identified resting-state (without DBS) neural biomarkers of relatively fast depression symptom variations within several days using multisite intracranial electroencephalography (iEEG) [44–46]. However, to date, identifying a neural biomarker that can simultaneously track the temporal dynamics of both slow and fast depression symptom variations, in particular during LHb DBS treatment, remains elusive.

Moreover, the neural biomarker needs to reflect the dose effect of different DBS parameters for optimizing stimulation parameters in aDBS. Since the DBS mechanism for treating TRD is largely unknown [47], only few studies have experimentally explored the dose effect of different DBS amplitudes on human neural signals [17,32,33]. On the other hand, DBS frequency also has been shown to play a key role in altering TRD symptoms [4–6,48]. However, how different LHb DBS parameters, especially stimulation frequencies, alter neural signals or neural biomarkers in TRD patients remains unknown.

Finally, the neural biomarker should possess a certain level of neurobiological interpretability. A critical neurobiological property that has been implicated in the pathophysiology of MDD [21–23] is the excitatory/inhibitory (E/I) balance of LHb activity. E/I balance refers to the equilibrium between synaptic excitation and inhibition received by neurons [49]. LHb has excitatory and inhibitory connections with a wide range of brain regions involved in the affect, reward, and cognition dimensions of MDD symptom, such as the dorsal raphe nucleus, nucleus accumbens, hippocampus, medial prefrontal cortex, etc [50]. Excessive excitation of LHb has been implicated in the pathology of MDD [21–23]. By contrast, electrical stimulation of LHb in animal depression models has been shown to inhibit overly-excited local neural activity, thus modulating the dopaminergic and serotoninergic activity and generate complex excitatory and inhibitory effects over a large network that may have led to depression-like behavior alleviation [51,52]. Furthermore, a prior study has found a correlation between pre-treatment LHb E/I balance with depression symptoms in DBS patients [34]. However, how LHb E/I balance changes during human patient DBS treatment and whether there exists a neural biomarker that can track such changes has not been investigated.

Here, to close the above gaps, we conducted LHb DBS on one TRD patient where we evaluated the patient’s symptoms and concurrently collected daily LHb LFP signals during the entire 41-week long treatment process (Figure 1). With this unique longitudinal dataset and by using machine learning techniques, we identified a clinically-viable neural biomarker from spectral and temporal LHb features, where the most contributing spectral and temporal features are the *β* (12-30 Hz) band power and Hurst exponent, respectively. Our identified neural biomarker (1) accurately classified high and low depression symptom severity states; (2) significantly tracked the temporal dynamics of weekly (slower) and daily (faster) depression symptom variations during the DBS treatment; (3) reflected the depression symptom changes in response to DBS frequency alterations. Finally, the neural biomarker can be interpreted as an indicator of the changes in LHb E/I balance during the DBS treatment. Together, our results have implications for identifying clinically-viable neural biomarkers to facilitate future LHb aDBS developments for treating TRD.

**Figure 1.**
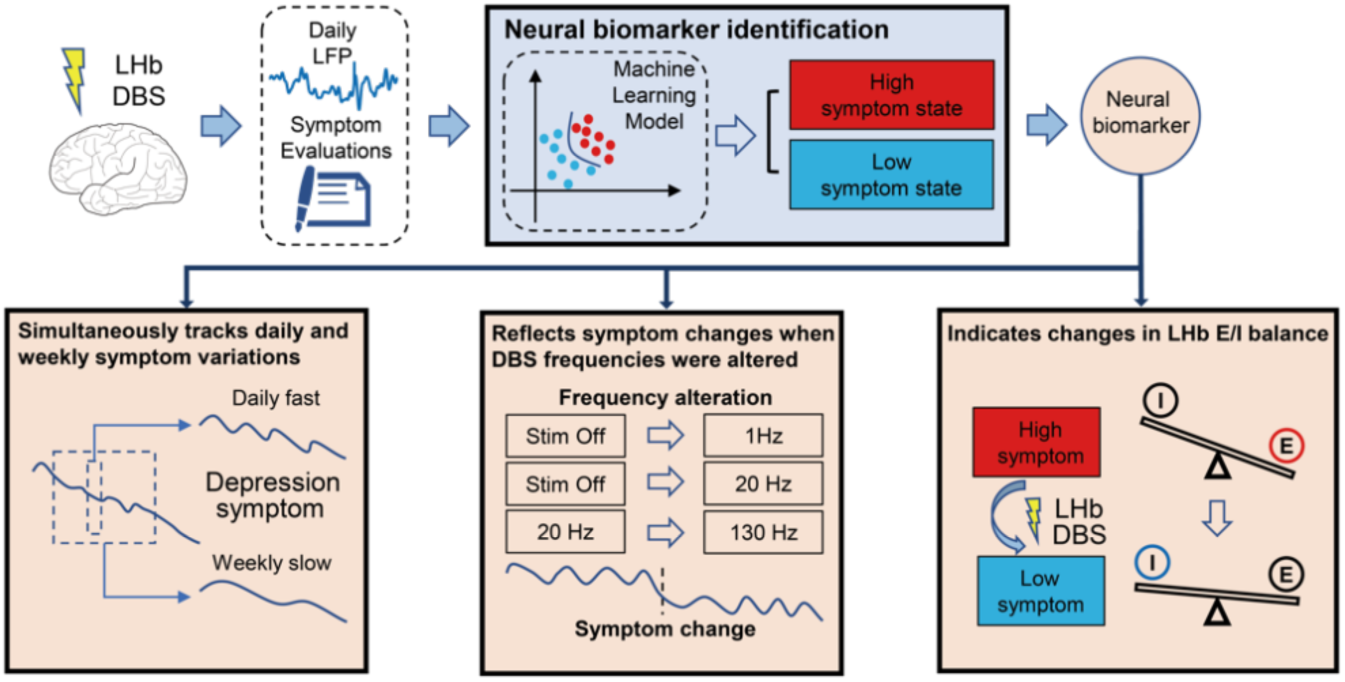
Study framework. During the LHb DBS treatment of one TRD patient, we used weekly clinical ratings and daily self-reports to evaluate the symptom variations, where we simultaneously collected daily LFP signals from LHb. Using machine learning models, we identified a neural biomarker that classified high and low depression symptom states during the DBS treatment. Using data not used in neural biomarker identification, we evaluated and interpreted the neural biomarker in terms of 1) simultaneously tracking the temporal dynamics of weekly slow and daily fast variations of depression symptoms; 2) reflecting symptom changes when DBS frequencies were altered; 3) indicating changes in LHb E/I balance.

## 2. Materials and Methods

### 2.1 Participant

This study included a male TRD patient aged 36-40 years old (see Note S1 for detailed patient medical information) participating in a clinical trial of LHb DBS treatment starting in October 2021. The patient provided informed consent for participation in the clinical trial. This study received approval from the Ethics Committee of Zhejiang University School of Medicine Second Affiliated Hospital (protocol number 20210218). It was registered at www.clinicaltrials.gov (IR2021001074), where detailed information regarding the inclusion and exclusion criteria can be accessed.

At the beginning of the clinical trial, two independent psychiatrists evaluated the patient’s psychotic symptoms using the 17-item Hamilton Depression Rating Scale (HAMD), the Montgomery Asberg Depression Scale (MADRS), and the Hamilton Anxiety Rating Scale (HAMA) as baseline assessments. In addition to the psychiatric assessments, the patient underwent a comprehensive physical examination, various mental scale assessments, and a magnetic resonance imaging (MRI) examination. We carefully ensured that other psychiatric diagnoses outlined in the Diagnostic and Statistical Manual of Mental Disorders-Fifth Edition (DSM-5) were excluded.

### 2.2 Surgical procedure

A standard DBS implantation procedure was employed. Bilateral quadripolar electrodes (1200-40, SceneRay, Suzhou, China) were surgically implanted in the LHb under local anesthesia (Figure 2A). The DBS electrodes had a diameter of 1.27 mm and a lead length of 400 mm. Each electrode’s four contacts measured 1.5 mm in length with a spacing of 0.5 mm. The LHb targeting was guided by preoperative MRI sequences. After confirming the absence of stimulation side effects through intraoperative testing, an implantable pulse generator (SR1101, SceneRay) was placed under general anesthesia. The DBS device was also capable of recording and wireless transmitting LFP signals (Figure 2B).

**Figure 2.**
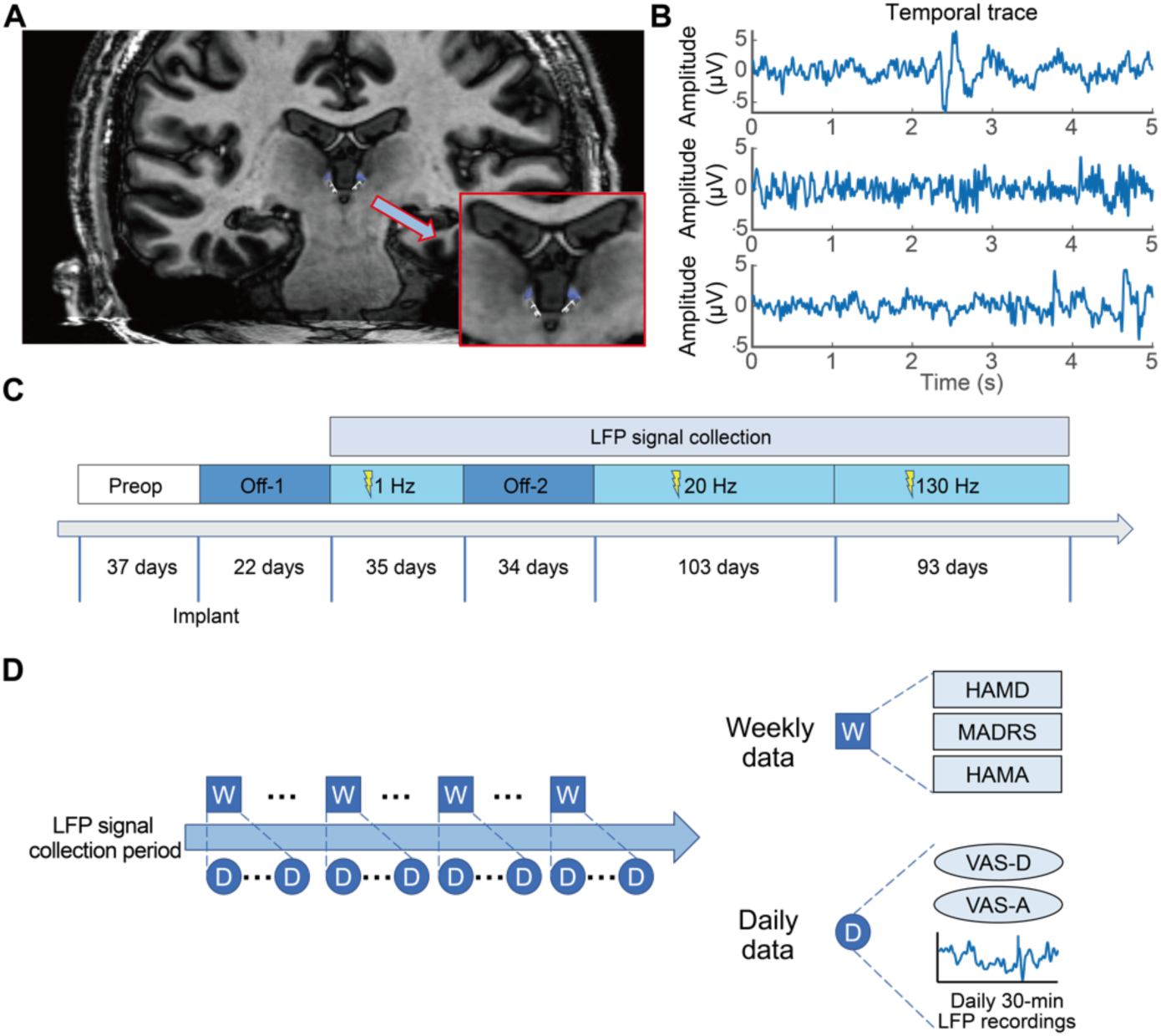
Experiment design. (A) MRI visualization showing the DBS lead placement within the patient’s LHb. The shaded blue area indicates the volume of tissue activated by the DBS. (B) Temporal dynamics of example epochs of LFP signals after preprocessing. (C) LHb DBS Treatment Timeline. The entire treatment process consisted of six stages. LFP signal collection began after the activation of 1 Hz stimulation. (D) Longitudinal data collection over time. Data were collected using two different time scales: 1) daily self-reports and 30-min LFP signals recorded around the self-reports; 2) weekly clinical ratings.

### 2.3 Fiber tracking of LHb

We used diffusion tensor images from the magnetic resonance imaging (MRI) examination to conduct fiber tracking of LHb with a standard procedure reported in a previous study [53]. First, we manually created the LHb as the region of interest (ROI) in standard Montreal Neurological Institute 152 (MNI152) space. The ROI in MNI152 standard space was then registered to the diffusion space of our patient for tractography. To ensure the exclusion of non-brain tissue, we employed the ’bet2’ function, and the cerebrospinal fluid mask was used as an exclusion mask. We further performed the probabilistic tractography processing using tools from the FSL Diffusion Toolbox. The LHb ROI was used as the seed mask to generate whole-brain distribution maps using FSL’s probtrackX tool. We performed 5000 streamline samples per voxel with distance normalization to ensure comprehensive coverage. Subsequently, the resulting fiber distribution maps were non-linearly registered back to the MNI152 standard space. To eliminate artifactual connections and noise, a threshold value of 1^7^ was implemented.

### 2.4 DBS treatment process and symptom evaluations

During the bilateral DBS treatment process, we made multiple adjustments to the stimulation parameters to achieve the best therapeutic effect. We divided the treatment process into six stages based on the alterations of stimulation parameters (Figure 2C): 1) the “Preop” stage, the time before DBS implantation; 2) the “Off-1” stage, patient recovery with DBS turned off; 3) the “1 Hz” stage, activation of 1 Hz stimulation; 4) the “Off-2” stage, DBS turned off because of unnoticed power off; 5) the “20 Hz” stage, re-activation of 20 Hz stimulation; 6) the “130 Hz” stage, activation of 130 Hz stimulation. More details can be found in Note S2. The entire duration of DBS treatment spanned 41 weeks (starting from DBS implantation).

The efficacy of DBS treatment was evaluated from two perspectives: clinician evaluation and self evaluation (Figure 2D). For clinician evaluation, a psychologist blinded to the current stimulation parameters and their adjustments evaluated the patient’s depression and anxiety symptoms on a weekly basis using standardized rating scales (HAMD, MADRS, HAMA). Response is defined as a 50^7^ or greater improvement on the HAMD score from the pre-treatment baseline. Remission is defined as achieving a HAMD score of 7 or less. The psychologist also evaluated the patient’s emotional blunting and cognitive functioning during the treatment. For every four to six months, the psychologist utilized the Oxford Depression Questionnaire (ODQ), a self-report tool to assess emotional blunting, along with two cognitive assessment tools (MATRICS Consensus Cognitive Battery (MCCB) and THINC-integrated tool (THINC-it)) to evaluate the patient’s cognitive abilities. For self evaluation, the patient used the Visual Analogue Scale (VAS) for depression (VAS-D) and anxiety (VAS-A) to self-report the symptom severity. Self-reported VASA and VAS-D had been used to assess the rapid effects of antidepressants [54]. To facilitate daily data collection, we established an online questionnaire system where the patient could conveniently complete the self-reports via the smartphone or computer.

### 2.5 LFP signal recording, signal processing, and feature extraction

After activating the 1 Hz stimulation, we collected daily LFP signals (30 minutes per day) concurrent with daily self-reported VAS-D and VAS-A (Figure 2D, details in section 2.4). LFP signals were recorded at a sampling rate of 1000 Hz. Notably, stimulation was deactivated during the signal acquisition process. We reconstructed the electrode positions using MRI and selected two contacts in the left hemisphere for bipolar recording of a single LFP channel. The patient was instructed to attempt daily LFP recording and VAS-D/VAS-A reporting. Throughout the entire 41-week (287-day) treatment, the patient was able to activate LFP recording and report VAS-D and VAS-A on 122 days distributed across 26 weeks. Therefore, the subsequent analyses focused on the LFP signals, VAS-D, and VAS-A scores recorded from these 122 days, and the HAMD, MADRS, and HAMA scores recorded for the 26 weeks.

Custom MATLAB scripts (MathWorks Inc., Natick, MA, USA) were used to preprocess the LFP signals. The LFP signals were first band-pass filtered from 1 to 30 Hz using a Butterworth filter of order 12 to avoid the noise observed in higher frequency bands. Then, we divided the daily 30-minute LFP signals into 10-second epochs with a 50^7^ overlap. Next, we used a standard procedure (details in Note S4) to remove bad epochs from daily LFP signals (example temporal traces of preprocessed LFP epochs were shown in Figure 2B).

For each remaining LFP epoch, we computed its spectral domain (SD) and temporal domain (TD) features. SD features included PSD of the four bands (*δ* (delta, 1-4 Hz), *θ* (theta, 4-8 Hz), *α* (alpha, 8-12 Hz), and *β* (beta, 12-30 Hz)) and phase-amplitude coupling (PAC) for six specific pairs of coupling. TD features included fourteen temporal domain features used in previous study [55], e.g., Hjorth mobility, Higuchi fractal dimension, Kurtosis, peak-to-peak amplitude, Hurst exponent, etc. These features capture the temporal properties of LFP from probabilistic distribution and information theory perspectives and have been widely used in brain signal analyses [56,57]. As a result, we obtained 24 features, comprising 10 SD features and 14 TD features for each LFP epoch. Details of these 24 features are included in Table S1 and Note S3. Finally, we averaged each feature across LFP epochs within the same day and obtained a single averaged 24-dimensional LFP feature vector. Our subsequent analyses were based on the daily LFP features as computed above.

### 2.6 Correlational analyses between LFP features and symptoms

We conducted Spearman’s rank correlation analyses between LFP features and symptoms. For each day, we correlated each daily LFP feature with the daily VAS-D and VAS-A. For each week, we computed the average of LFP features across the days that belonged to this week, resulting in weekly LFP features; we then correlated each weekly LFP feature with the weekly clinical evaluation scales HAMD, HAMA, and MADRS. Bonferroni correction was used to adjust for multiple comparisons.

### 2.7 Identification of neural biomarker

Next, we used a data-driven method to identify an LHb neural biomarker of depression symptoms, where we built a machine learning model to use LFP features to classify high and low depression symptom states.

First, we defined the high and low depression symptom states of the patient by k-means clustering the weekly depression scales HAMD and MADRS similar to prior work [17]. Among the total 26 weeks (122 days) of LFP data, 7 weeks (29 days) of LFP data belonged to the low depression symptom state (labeled 0), 4 weeks (22 days) of LFP data belonged to the high depression symptom state (labeled 1). The remaining 15 weeks (71 days) belonging to the transition symptom state were unlabeled and used as test data for subsequent biomarker tracking evaluation (see next section).

Second, based on the labeled data, we built a machine learning model to use the LFP features to classify high and low depression symptom states. We constructed six machine learning models: logistic regression (LR), multilayer perceptron, adaptive boosting, support vector machine, random forest, and linear discriminant analysis. We trained and tested these models using 5-fold cross-validations that were repeated 200 times, where we computed the averaged cross-validated classification accuracy, specificity, sensitivity, F1 score, and Receiver Operating Characteristic (ROC) Area Under the Curve (AUC) score as the performance metrics. The model with the highest accuracy was selected for further analyses.

Third, the chosen model was retrained with all labeled data, leading to a “neural biomarker model”. This model takes the LFP feature as input and outputs the decision variable as the neural biomarker value (e.g., in the LR model, the decision variable was computed from the decision probability via the inverse sigmoid function). This allows us to compute a neural biomarker value for any given LFP feature. Higher neural biomarker values indicate more severe depression symptoms.

In essence, our identified neural biomarker aggregates spectral and temporal domain features from the LHb LFP signal to classify high and low depression states during DBS treatment.

### 2.8 Evaluation of the neural biomarker

We evaluated the neural biomarker in terms of (1) tracking the temporal dynamics of weekly symptom variations; (2) tracking the dynamics of daily symptom variations; (3) reflecting changes in symptom variations induced by DBS frequency alterations.

First, we investigated tracking the temporal dynamics of weekly depression and anxiety symptom scales that were not used in neural biomarker identification. We took the daily LFP features as inputs to the neural biomarker model and computed the output daily neural biomarkers. We then averaged the daily neural biomarkers belonging to the same week to compute the weekly neural biomarkers. We next correlated the weekly neural biomarker values with the weekly HAMD, MADRS, and HAMA scores, respectively, using Spearman’s rank correlation analyses with explained variance (EV) as an estimation. We further analyzed the temporal dynamics in the neural biomarker and symptoms, using the dynamic time warping (DTW) distance [58] to measure the temporal tracking ability of the neural biomarker. Both the neural biomarker values and the symptom scales were normalized to a range of 0 to 1. We used a size three Sakoe–Chiba warping window in the DTW analysis following prior work [17,59]. A smaller DTW distance represents better temporal tracking. To determine the significance of the computed DTW distance, we randomly shuffled the temporal sequence of the neural biomarker 10,000 times and used the corresponding shuffled DTW distances as the null hypothesis distribution for computing the *P* value.

Second, we investigated tracking the temporal dynamics of the daily VAS-D and VAS-A self-reports, which were also not used in neural biomarker identification. Similar to the weekly case, daily LFP features were used to generate daily neural biomarkers, which were then correlated with daily VAS-D and VAS-A reports. DTW was again used to assess the temporal tracking of daily depression symptom variations.

Third, we qualitatively compared trends in weekly neural biomarkers and depression ratings across three DBS frequency alterations (1 Hz to Off-2, Off-2 to 20 Hz, 20 Hz to 130 Hz). We used the two-sided Wilcoxon rank-sum test to check whether there was a significant difference between the two stages before and after alteration. We also averaged the neural biomarker values and depression ratings across five time periods for each case: 1) from the beginning of this stage to two weeks before the alteration week; 2) during the week before the alteration week; 3) during the alteration week; 4) during one week after the alteration week; 5) averaged from two weeks after the alteration week to the end of this stage.

### 2.9 Interpretation of the neural biomarker

We then conducted multiple analyses to interpret the neurobiological implications of the identified neural biomarker. First, we recognized the most contributing spectral and temporal LFP features of the neural biomarker. To evaluate their contributions, we performed individual classification of the high and low symptom states using each feature separately. The performance of the classification provided an indication of the level of information contained within each feature regarding the symptom state. To further validate the contribution of these features, we compared the weights of each feature in the neural biomarker identification model (see Section 2.7).

Second, we related the neutral biomarker to the LHb E/I balance. Following prior work [60], we computed the LFP spectrum’s 1/*f* slope as a quantitative indicator of E/I balance, where a larger absolute 1/*f* slope indicates more inhibition. More specifically, after calculating the PSD of the remaining epochs after the removal of bad epochs (see Section 2.5), we averaged the spectrum across LFP epochs within the same day and selected a frequency range of interest to be 1-30 Hz (our signal spectrum range). We then estimated 1/*f* slope from the log-transformed PSD using linear regression, with the coefficient representing the 1/*f* slope. We next normalized the E/I indicator by computing the percentage change relative to the 1/*f* slope of the first LFP recording day. We finally correlated this E/I indicator with the identified neural biomarker and its most contributing features using standard Spearman’s rank correlation analyses.

## 3. Results

### 3.1 LHb DBS improved the patient’s depression symptoms, emotional blunting, and cognitive functions

We first examined the TRD patient’s depression and anxiety symptom changes throughout the LHb DBS treatment process. At the beginning of treatment, the patient’s baseline HAMD score was 20, MADRS score was 25, and HAMA score was 16. In terms of the weekly clinical ratings (Table 1 and Figure 3A), the patient responded at week 14 (HAMD score dropped to 10; MADRS score dropped to 19; HAMA score dropped to 8) and achieved remission by the end of the 41-week treatment (HAMD score was 7; MADRS score was 9; HAMA score was 6). The daily self-reports followed a similar decreasing trend (Figure 3B). Such a consistent trend was confirmed by the strong positive correlation between the daily self-reports and weekly clinical ratings (Spearman’s *ρ* > 0.5 , *P* < 0.05 for all pair-wise correlations; see Table S2 and Figure S1 for details).

**Figure 3.**
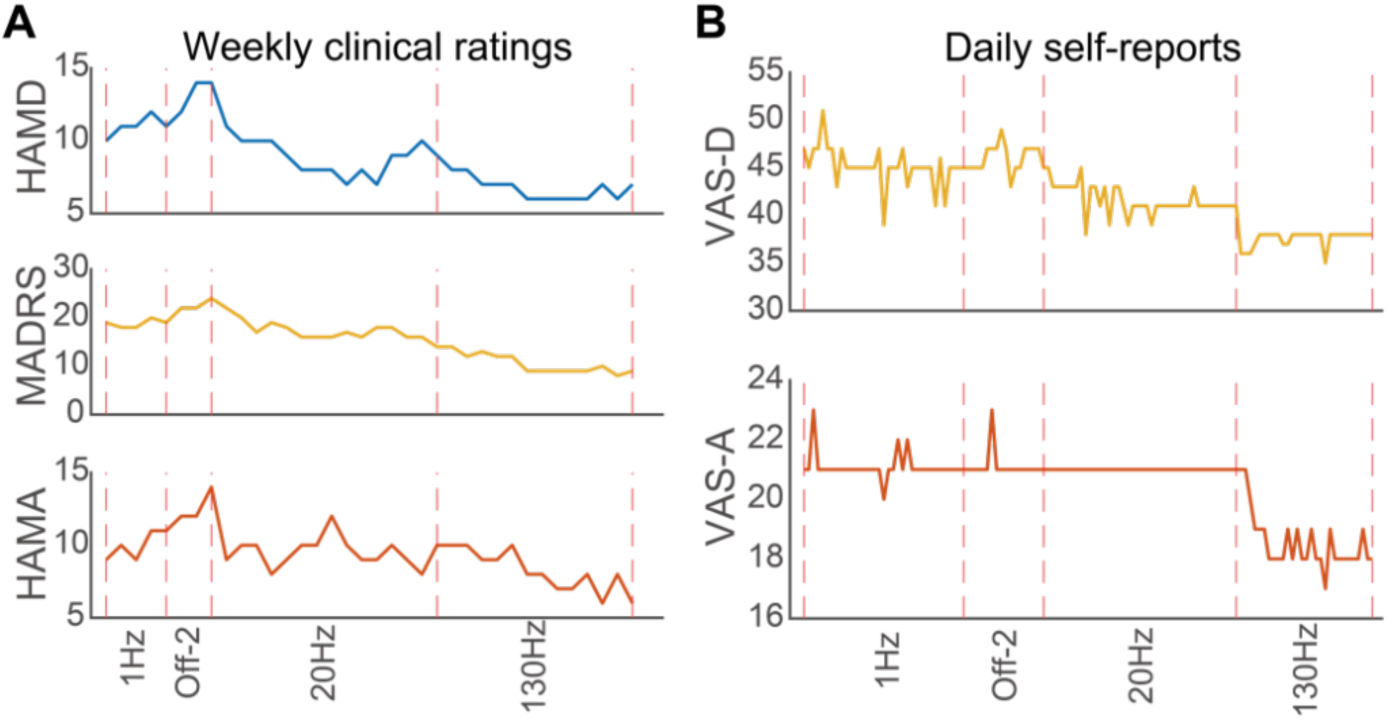
Changes of weekly and daily symptom scores during the LHb DBS treatment. (A) Changes of weekly clinical ratings during the treatment. The vertical dashed lines represent different treatment stages indicated by the x-axis labels. (B) Changes of daily self-reports during the treatment. The vertical dashed lines represent different treatment stages indicated by the x-axis labels.

**Table 1.** Assessment of depression and anxiety symptoms with weekly clinical ratings. mean±s.e.m.

| Weekly clinical ratings | LHb DBS treatment stage |  |  |  |  |
| --- | --- | --- | --- | --- | --- |
|  | Baseline<br>N=1 | 1Hz<br>N=4 | Off-2<br>N=3 | 20Hz<br>N=15 | 130Hz<br>N=14 |
| **HAMD** | 20 | 11.00±0.41 | 12.33±0.88 | 9.20±0.46 | 6.86±0.25 |
| **MADRS** | 25 | 18.75±0.48 | 21.00±1.00 | 17.93±0.63 | 10.64±0.56 |
| **HAMA** | 16 | 9.75±0.48 | 11.67±0.33 | 9.80±0.39 | 8.29±0.38 |

Besides alleviating depression and anxiety symptoms as indicated by the weekly clinical ratings and daily self-reports, LHb DBS also improved the patient’s emotional blunting and cognitive functioning. The total scores of ODQ dropped from the presurgery baseline of 117 to 94, and most of the ODQ subdomains continuously decreased during the DBS treatment (Table 2), indicating an improvement of emotional blunting. MCCB and THINC-it as cognitive function instruments suggested cognitive performance improved compared to the presurgery baseline (Table 3). Specifically, in the MCCB test, persistent improvements were found in processing speed, verbal learning, visual learning, reasoning/problem solving, and social cognition, while attention and working memory temporarily improved but fluctuated during the treatment. In the THINC-it test, the performance of Symbol check and Trail task, which were highly related to working memory [61] and executive functioning [62], showed persistent improvements, while PHQ-D-5, Spotter, and CodeBreaker task performance fluctuated.

**Table 2.** Assessment of emotional blunting with Oxford Depressive Symptoms Questionnaire (ODQ).

| Domains of ODQ | Follow up after activation of stimulation |  |  |  |
| --- | --- | --- | --- | --- |
|  | Baseline | 69 days | 173 days | 270 days |
| General Reduction | 23 | 24 | 23 | 19 |
| Reduction in Positive | 25 | 25 | 24 | 20 |
| Emotional Detachment | 25 | 25 | 16 | 18 |
| Not Caring | 22 | 23 | 18 | 18 |
| Antidepressant as Cause | 22 | 22 | 18 | 19 |
| Total | 117 | 119 | 99 | 94 |

**Table 3.**
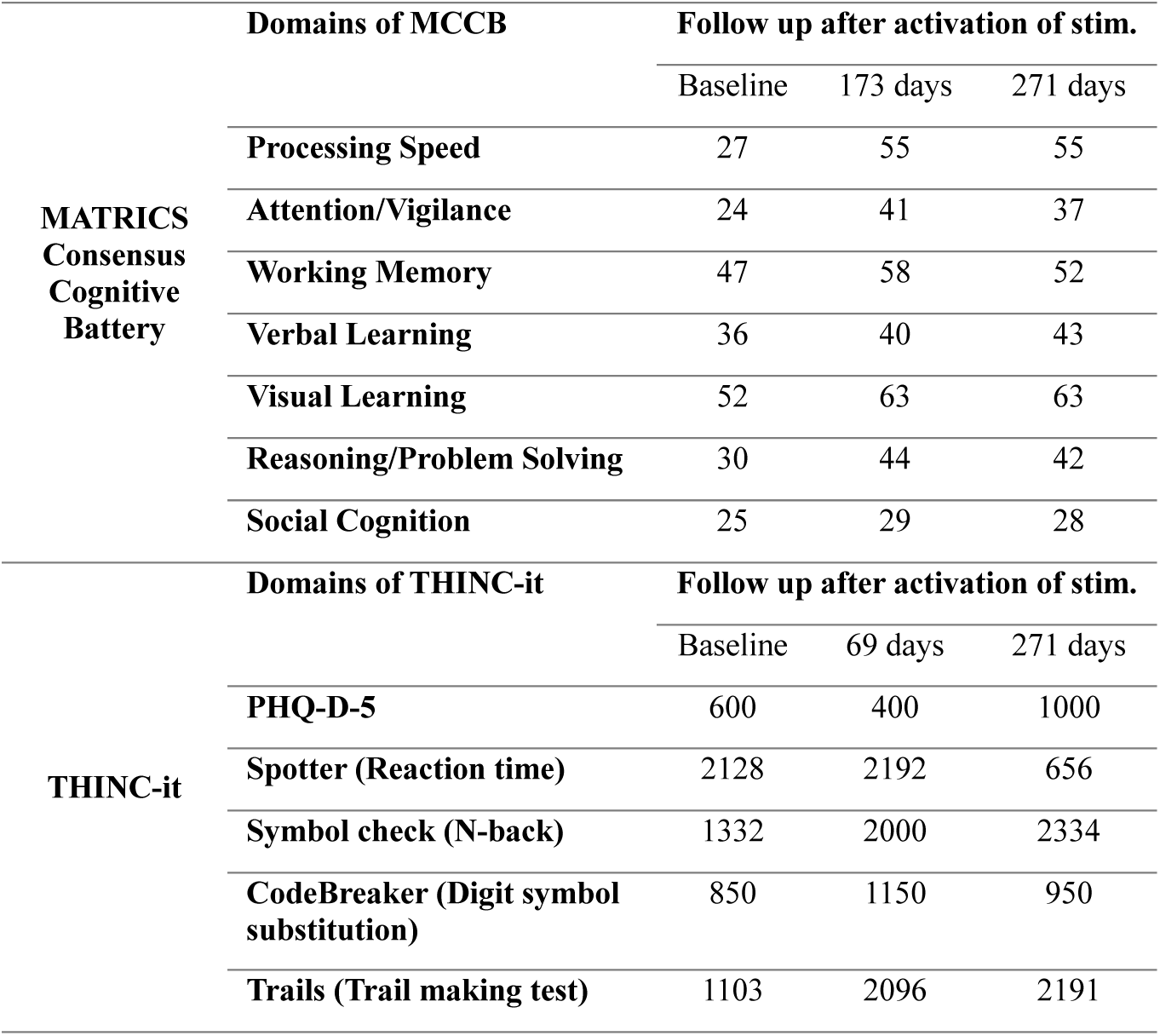
Assessment of cognitive functioning with MATRICS Consensus Cognitive Battery (MCCB) and THINC-it.

### 3.2 Multiple spectral and temporal LHb LFP features significantly correlated with symptom changes

During the LHb DBS treatment process, we recorded daily LHb LFP signals. Therefore, we then investigated how the LFP features correlated with the patient’s symptom changes (Figure 4A). We found that many of the temporal and spectral domain LHb LFP features were significantly correlated with the weekly clinical ratings and daily self-reports (Figure 4B). For example, Hurst exponent exhibited the strongest correlations with both weekly HAMD scores and daily VAS-D scores (Figure 4C). The results indicate that it is feasible to identify an LHb neural biomarker of depression symptoms from the LFP temporal and spectral domain features.

**Figure 4.**
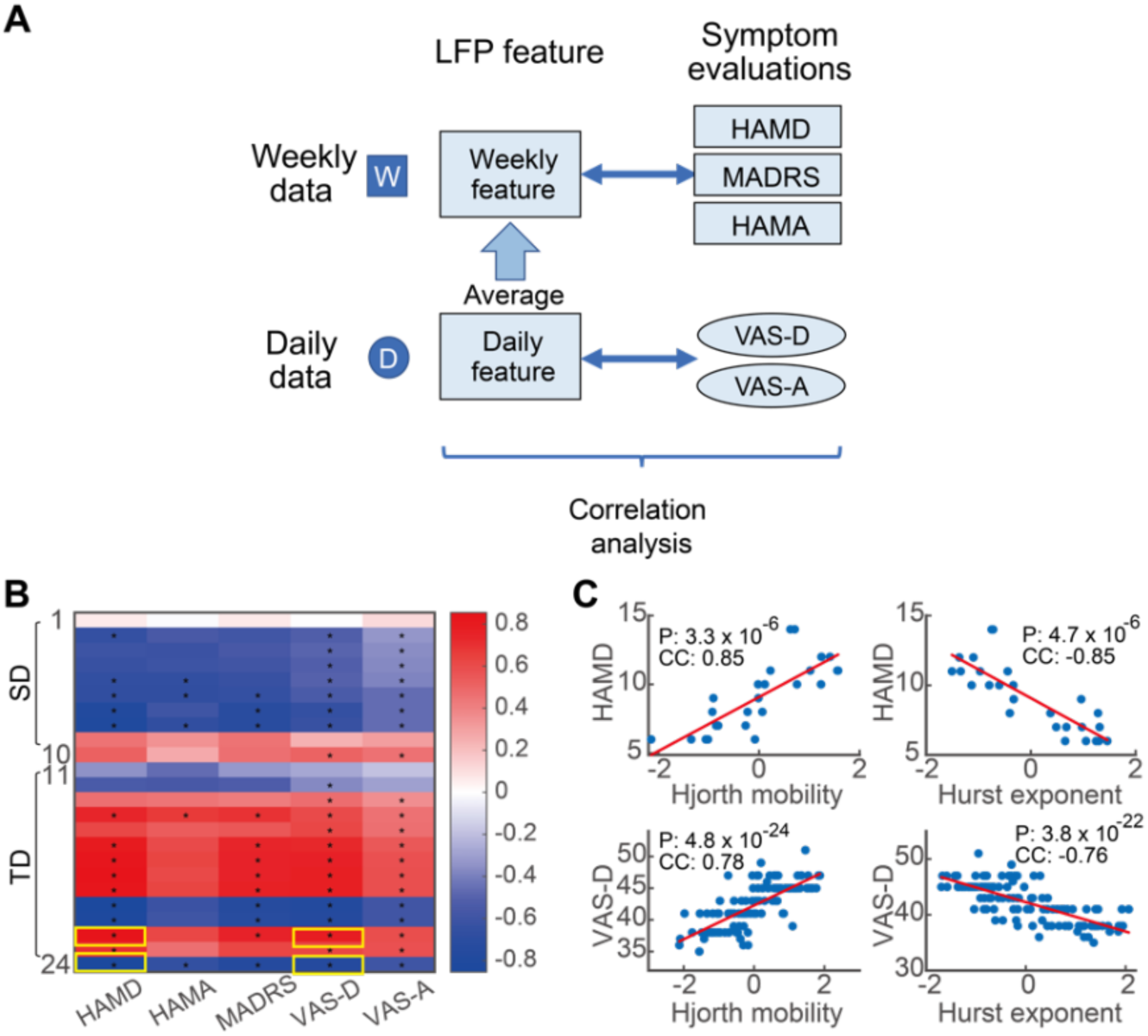
Correlations between the symptom evaluations (weekly and daily symptom scores) and the LFP features (temporal and spectral domain features). (A) Schematic diagram of correlation analysis. (B) Heatmap of the correlation coefficients. Each cell shows the correlation coefficient (CC) value between one LFP feature (y-axis) and one symptom score (x-axis), and cells marked with * indicate the coefficients that are significantly different from zero (Bonferroni corrected *p*<0.05). (C) Positive and negative correlation examples with the weekly HAMD score and daily VAS-D score as shown by the yellow boxes in (B).

### 3.3 Accurate classification of high and low symptom severity states led to the identification of an LHb neural biomarker

We next used the LFP temporal and spectral features to identify a neural biomarker that can classify high and low depression symptom severity states (Figure 5A). We started by defining a state of high symptom severity and a state of low symptom severity via clustering the weekly depression scales HAMD and MADRS (Figure 5B). The high symptom state (7 weeks) had an average HAMD score of 12.8 and an average MADRS score of 22.5, while the low symptom state (4 weeks) had an average HAMD score of 6.3 and an average MADRS score of 9.0. We then used LFP temporal domain and spectral domain features from these 11 weeks to classify the high and low symptom severity states via six machine learning models in cross-validation. Among these six models, the LR model performed better than other more complicated models (Figure 5C and table S3). Specifically, for the LR model, the cross-validated classification accuracy was 0.973±0.002 (mean ± s.e.m.), the specificity was 0.961±0.003, the sensitivity was 0.988±0.002, the F1-score was 0.970±0.002, and the AUC score was 0.974±0.001, which were all significantly higher than other models (Wilcoxon rank sum test, Bonferroni corrected *P* < 0.05 for all comparisons), suggesting that the LR model was best suited for classifying the collected data. We thus selected the LR model for further analyses. Then, we retrained the LR model using the 11 weeks’ labeled data, resulting in the neural biomarker model, which model took the LFP features as input and output the model decision value as the neural biomarker, with higher values indicating worse symptoms.

**Figure 5.**
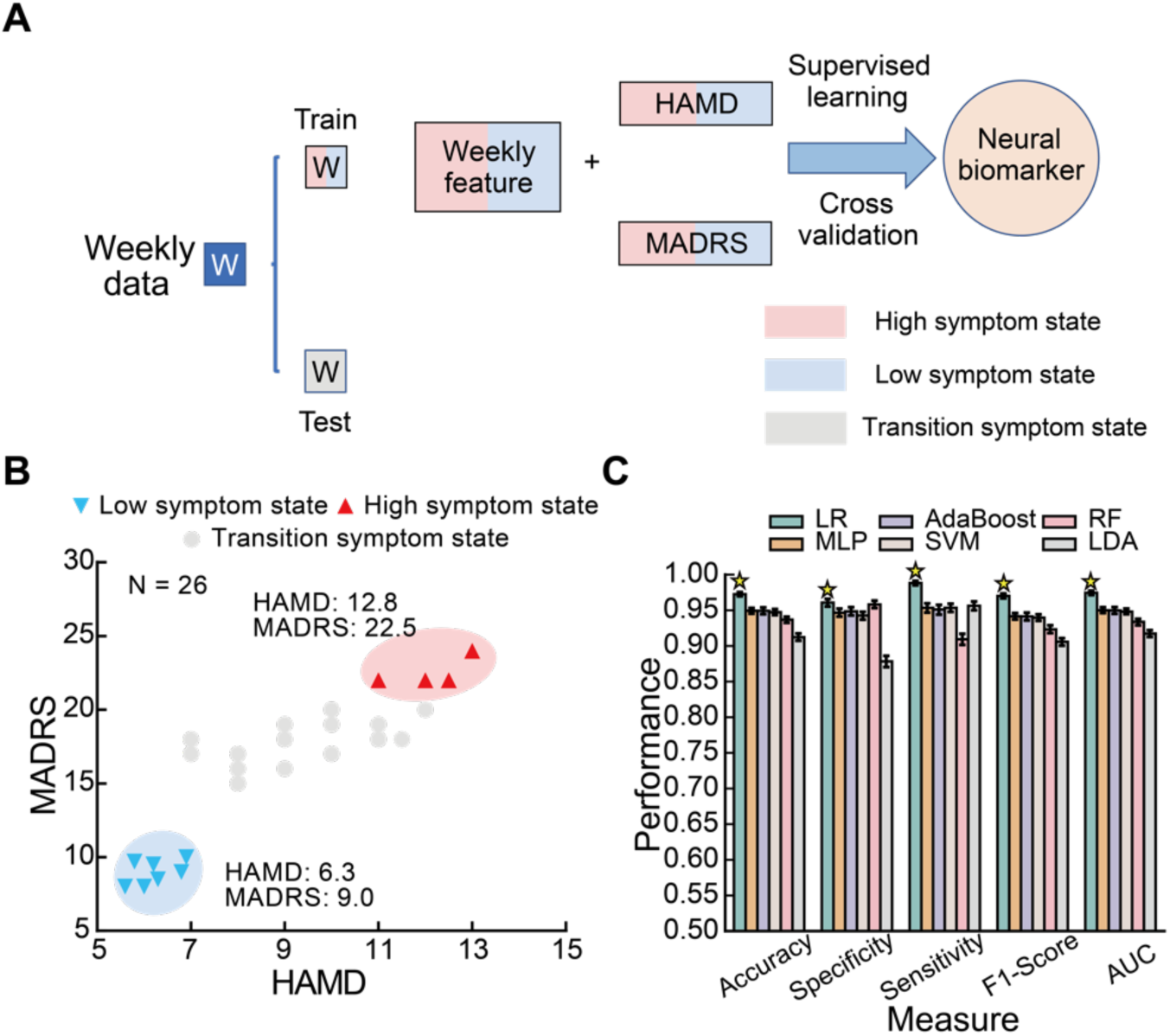
Accurate classification of high and low depression symptom severity states by the identified neural biomarker. (A) Schematic diagram of neural biomarker identification. Note that the identification was conducted using supervised learning and cross-validation purely based on the weekly data in the high and low symptom states. The rest weekly data in the transition symptom state were used later to test the tracking performance of the identified neural biomarker. (B) Clustering of depression symptom severity states. We clustered the HAMD and MADRS scores to obtain three distinct symptom states: a high depression symptom severity state (shaded in red), a low depression symptom severity state (shaded in blue), and a transition state (shaded in grey). Each point represents data of a week. The average HAMD and MADRS scores of the high and low clusters are also indicated in the figure. (C) Classification performance of different classifiers. The bar represents mean and the whiskers represent the 95% confidence interval. Six classification models (different colors) were compared in terms of five performance matrices (different x-axis groups): accuracy, specificity, sensitivity, F1-Score and AUC. The best model was indicated by a yellow star for each metric. Classification model abbreviations: logistic regression (LR), multilayer perceptron (MLP), adaptive boosting (AdaBoost), support vector machine (SVM), random forest (RF), and linear discriminant analysis (LDA).

### 3.4 The identified neural biomarker simultaneously tracked the temporal dynamics of weekly and daily depression symptom variations during LHb DBS treatment

After identifying the neural biomarker of depression symptoms, we evaluated its ability to track the temporal dynamics of slow (weekly) and fast (daily) depression symptom variations during the LHb DBS treatment. For slow weekly variations, we used the identified neural biomarker model to compute weekly neural biomarker values (see Section 2.8). We used the weekly neural biomarker values to predict the associated weekly clinical ratings, where we strictly excluded the weekly data that were used to identify the neural biomarker (i.e., the prediction was based on unseen transition symptom state data not used in training the neural biomarker model, Figure 6A). We found that the weekly neural biomarker values significantly predicted the HAMD scores (Figure 6B, EV=0.74, *P* = 1.1 × 10^-“^). Further, considering the temporal dynamics in detail by using the DTW distance analysis (see Section 2.8), we found that the weekly neural biomarker significantly tracked the temporal dynamics of weekly HAMD score variations (random shuffle *P* = 0.00001). Consistently, the weekly neural biomarker values significantly predicted the MADRS scores (Figure 6C, EV=0.34, *P* = 0.039), and tracked the temporal dynamics in the DTW distance analysis with marginally significant statistics (random shuffle *P* = 0.1079). Conversely, the weekly neural biomarker values did not predict the HAMA scores (Figure 6D, EV=0.03, *P* = 5.2 × 10^-1^) or tracked the temporal dynamics (random shuffle *P* = 0.5865).

**Figure 6.**
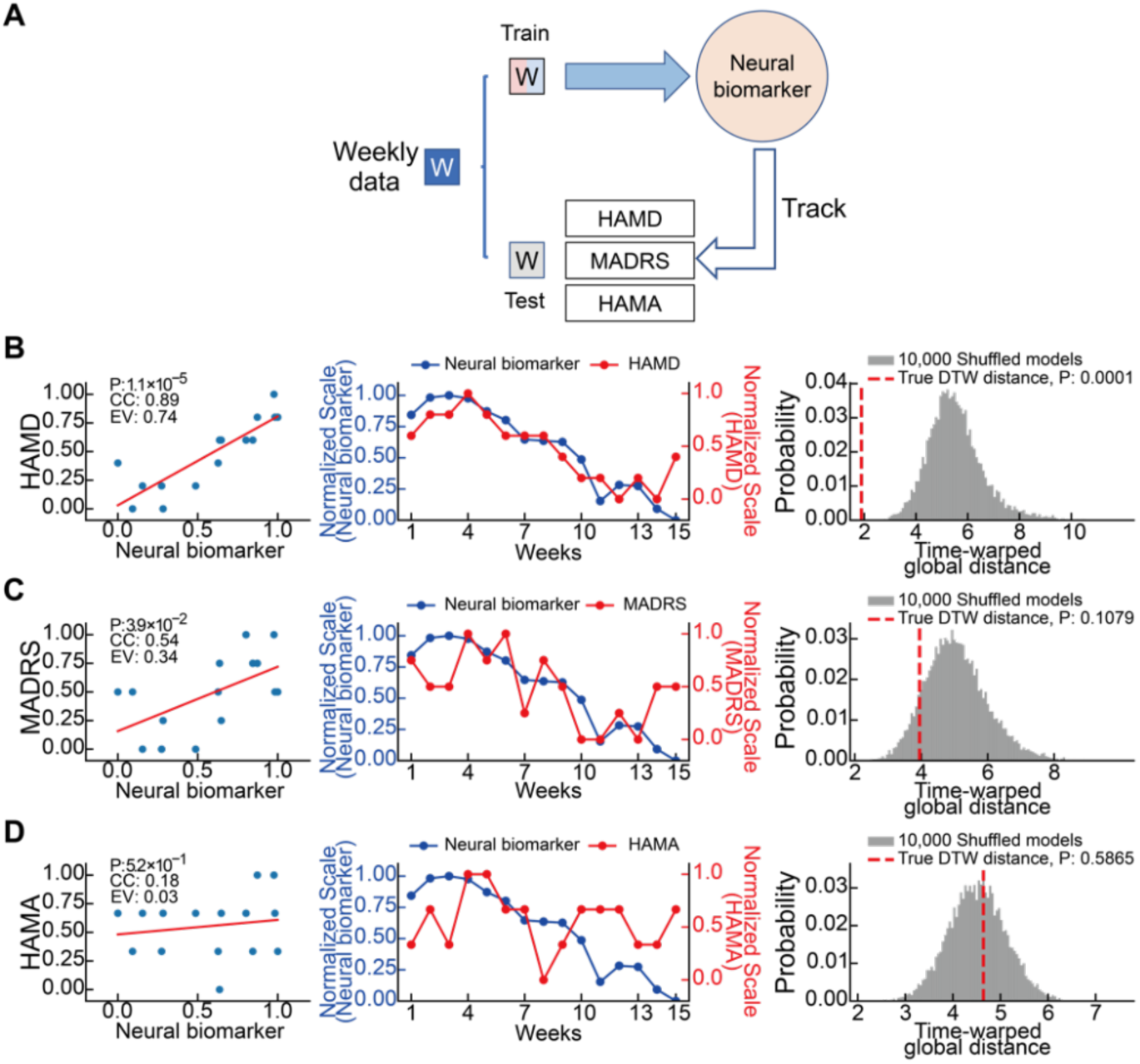
Accurate tracking of the temporal dynamics of weekly depression symptom variations by the identified neural biomarker. (A) Schematic diagram of analyzing the neural biomarker’s weekly tracking performance. The neural biomarker, derived from the training data of weekly high and low symptom states, was tested by tracking the unseen weekly clinical ratings of transition symptom state. (B) Left: correlation between the identified neural biomarker values and the weekly HAMD scores. CC: correlation coefficient. EV: Explained variance. Middle: neural biomarker tracking of the weekly HAMD score dynamics over time. Right: the DTW distance analysis result for evaluating the significance of tracking in the middle panel. Smaller DTW distance represents better tracking. Note that we normalized both the neural biomarker values and the symptom scales to a range of 0 to 1 using min-max normalization for better visualization. (C) same as (B) but for the weekly MADRS scores. (D) same as (B) but for the weekly HAMA scores.

For fast daily variations, we used the identified neural biomarker model to compute daily neural biomarker values and used the daily neural biomarker values to predict the associated daily self-reports (again, data not used in training the neural biomarker model, Figure 7A). We found that the daily neural biomarker values significantly predicted the VAS-D scores (Figure 7B, EV=0.63, *P* = 1.3 × 10^-27^) and showed significant tracking of VAS-D dynamics (random shuffle *P* = 0.0001). By contrast, while the daily neural biomarker values predicted the VAS-A scores (Figure 7C, EV=0.51, *P* = 5.3 × 10^-16^) but the daily neural biomarker did not significantly track VAS-A dynamics (random shuffle *P* = 1.00).

**Figure 7.**
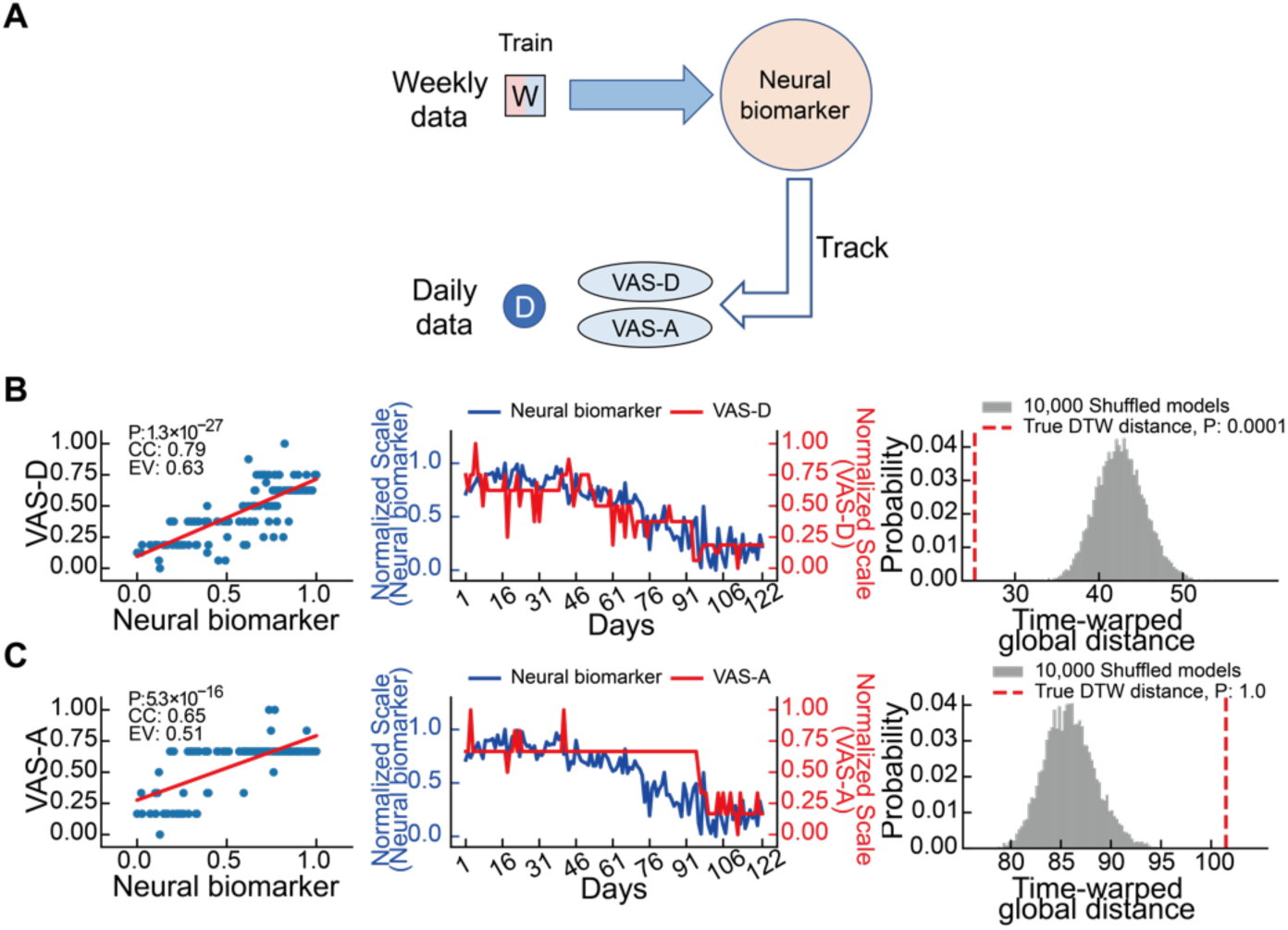
Accurate tracking of the temporal dynamics of daily depression symptom variations by the identified neural biomarker. (A) Schematic diagram of analyzing the neural biomarker’s daily tracking performance. The neural biomarker, derived from the training data of weekly high and low symptom states, was tested by tracking the unseen daily self-reports. (B) Left: correlation between the identified neural biomarker values and the daily VAS-D scores unseen in neural biomarker identification. EV: Explained variance. Middle: neural biomarker tracking of the daily VAS-D score dynamics over time. Right: the DTW distance analysis result for evaluating the significance of tracking in the middle panel. Smaller DTW distance represents better tracking. (C) same as (B) but for the daily VAS-A scores.

In summary, the results show that the identified neural biomarker significantly tracked the temporal dynamics of both weekly and daily variations in depression symptoms during the LHb DBS treatment and specifically tracked depression symptoms rather than anxiety symptoms.

### 3.5 The identified neural biomarker reflected changes of depression symptoms in response to DBS parameter alterations

A useful neural biomarker for DBS also needs to reflect the effect of different DBS parameters. We thus evaluated if the identified neural biomarker could reflect changes in depression symptoms in response to DBS parameter alterations. We applied three different DBS frequencies during the treatment: 1 Hz, 20 Hz, and 130 Hz. For the DBS alteration from 1 Hz to stimulation off (Off-2, the DBS device shut down due to unnoticed power off), there was a trend of increasing for the neural biomarker, HAMD, and MADRS while the statistical tests were not significant due to the limited sample size (Figure 8A, 1Hz vs. stimulation off, normalized mean±s.e.m., neural biomarker: 0.912±0.055 vs. 0.956±0.026, *P*=0.35; HAMD: 0.562±0.062 vs. 0.688±0.036, *P*=0.16; MADRS: 0.656 ± 0.031 vs. 0.734 ± 0.053, *P*=0.35), which indicated a rebound trend of depression symptoms due to the disruption of DBS treatment. For the DBS alteration from stimulation off (Off-2) to 20 Hz stimulation, the neural biomarker, HAMD, and MADRS consistently decreased (Figure 8B, stimulation off vs. 20 Hz, neural biomarker: 0.905 ± 0.095 vs. 0.545 ± 0.059, *P*=0.04; HAMD: 0.875±0.125 vs. 0.375±0.072, *P*=0.05; MADRS: 0.875±0.000 vs. 0.606±0.061, *P*=0.08). Specifically, the neural biomarker, HAMD and MADRS scores all decreased at the week of DBS frequency alteration, further decreased one week after the alteration and continued to decrease with more obvious changes after week two. For the DBS alteration from 20 Hz to 130 Hz stimulation, the neural biomarker, HAMD, and MADRS also consistently decreased (Figure 8C, 20 Hz vs. 130 Hz, neural biomarker: 0.524±0.060 vs. 0.088±0.029, *P*=0.001; HAMD: 0.323±0.054 vs. 0.042±0.026, *P*=0.004; MADRS: 0.573±0.056 vs. 0.062±0.016, *P*=0.001). More specifically, the neural biomarker, HAMD and MADRS scores already showed a trend of decreasing before the DBS frequency alteration, and the alleviated symptoms stayed relatively stable during the alteration week, at week one after the alteration, and the same stable trend continued after week two. The results suggested that the 1 Hz DBS did not induce an obvious change in depression symptoms, while the 20 Hz and 130 Hz DBS had more meaningful effects. The results further showed that the neural biomarker indeed reflected the different change patterns in depression symptoms when the DBS frequencies were altered.

**Figure 8.**
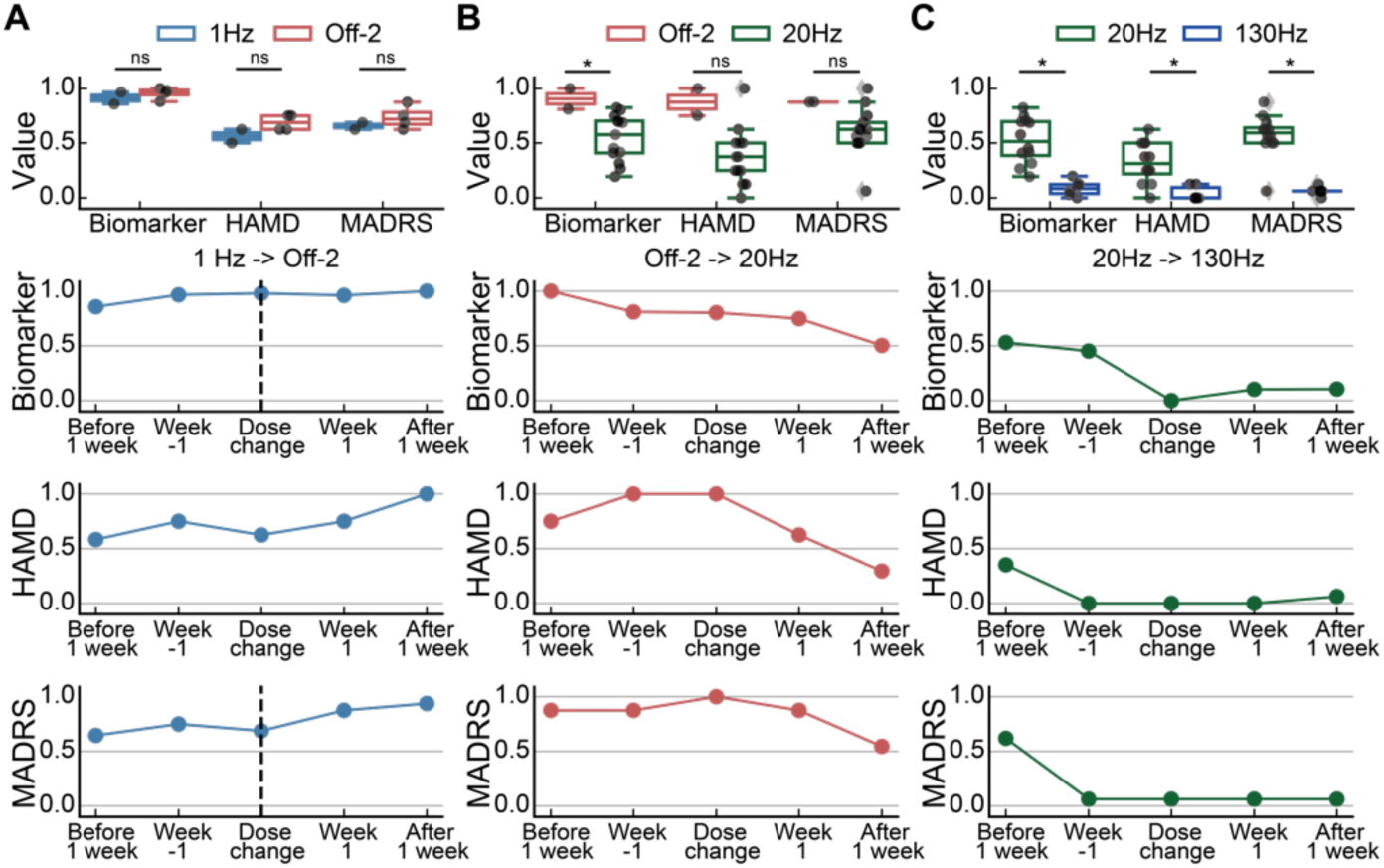
The consistent trend between the neural biomarker and the depression symptom changes when DBS frequencies were altered. (A) DBS frequency was altered from 1 Hz to Off-2. Top: comparison of the values of the biomarker, HAMD scores and MADRS scores between the 1Hz stage and Off-2 stage. Two-sided Wilcoxon rank-sum test was used for significance test. Asterisks indicate the significance \**p*<0.05 and ns indicates no significance. Row two: changes in the identified neural biomarker time-locked to the DBS frequency alteration week (vertical dashed line). Row three: changes in HAMD time-locked to the DBS frequency alteration week. Bottom: changes in MADRS time-locked to the DBS frequency alteration week. (B) same as (A) but for DBS frequency alteration from Off-2 to 20 Hz. (C) same as (A) but for DBS frequency alteration from 20 Hz to 130 Hz.

### 3.6 Neural biomarker identification and testing was robust to adequate decreasing of data sample size

The neural biomarker was identified and tested by a dataset consisting of 122 days of data samples. Due to the difficulty of obtaining a large amount of longitudinal data, we investigated the robustness of neural biomarker identification and testing in terms of data sample size decreasing. We decreased the data sample size gradually from 122 to 10 with a step size of 2, randomly removed data from neural biomarker identification and testing for each data sample size, and then repeated the entire neural biomarker classification and tracking analyses. We found that the neural biomarker classification accuracy consistently maintained above 0.95 even by decreasing the sample size to 70 but had a large decrease below a sample size of 40 (Figure 9A). Similarly, the neural biomarker tracking performance (EV in prediction) maintained relatively stable by decreasing the sample size to 70 but had much larger fluctuations below a sample size of 40 (Figure 9B). These results suggest that our data sample size of 122 days was sufficient to obtain a robust neural biomarker.

**Figure 9.**
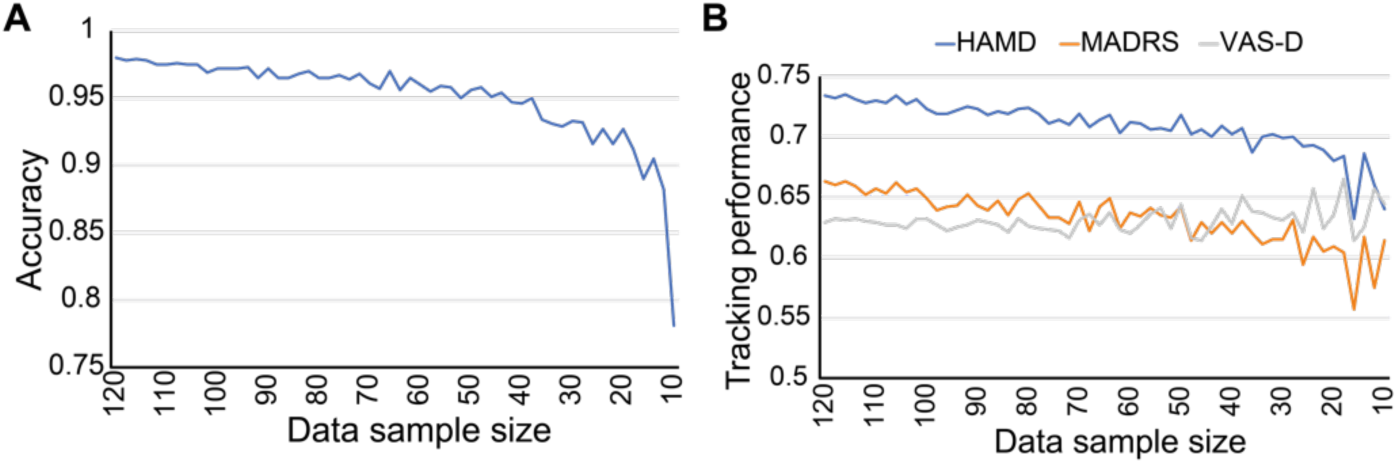
Changes of the neural biomarker identification and tracking performance by gradually decreasing the data sample size. (A) The accuracy of classifying the high and low symptom states. (B) The tracking performance of depression symptoms (weekly HAMD, weekly MADRS, and daily VAS-D).

### 3.7 The identified neural biomarker was mainly contributed by the *β* band spectral feature and the Hurst exponent temporal feature

Having demonstrated the usefulness and robustness of the identified neural biomarker, we next aimed to provide neurobiological interpretations for the neural biomarker from multiple aspects. We started by investigating the most contributing spectral and temporal LFP features in the neural biomarker. We recognized the most contributing spectral and temporal LFP features by separately classifying the high and low weekly symptom states using each individual spectral and temporal LFP feature (Figure 10A). The best temporal domain feature was the Hurst exponent, with a cross-validated classification accuracy of 0.93. While the best spectral domain feature was the *β* band PSD, with an average accuracy of 0.80. Overall, temporal domain features (average accuracy: 0.78) outperformed spectral domain features (average accuracy: 0.71). Notably, combining all the LFP features yielded superior performance compared to individual features. Consistently, by investigating the logistic regression coefficients of the neural biomarker model (Figure 10B), we found that the features with better classification accuracy also had larger coefficients in the neural biomarker model, again showing the importance of *β* band PSD and Hurst exponent.

**Figure 10.**
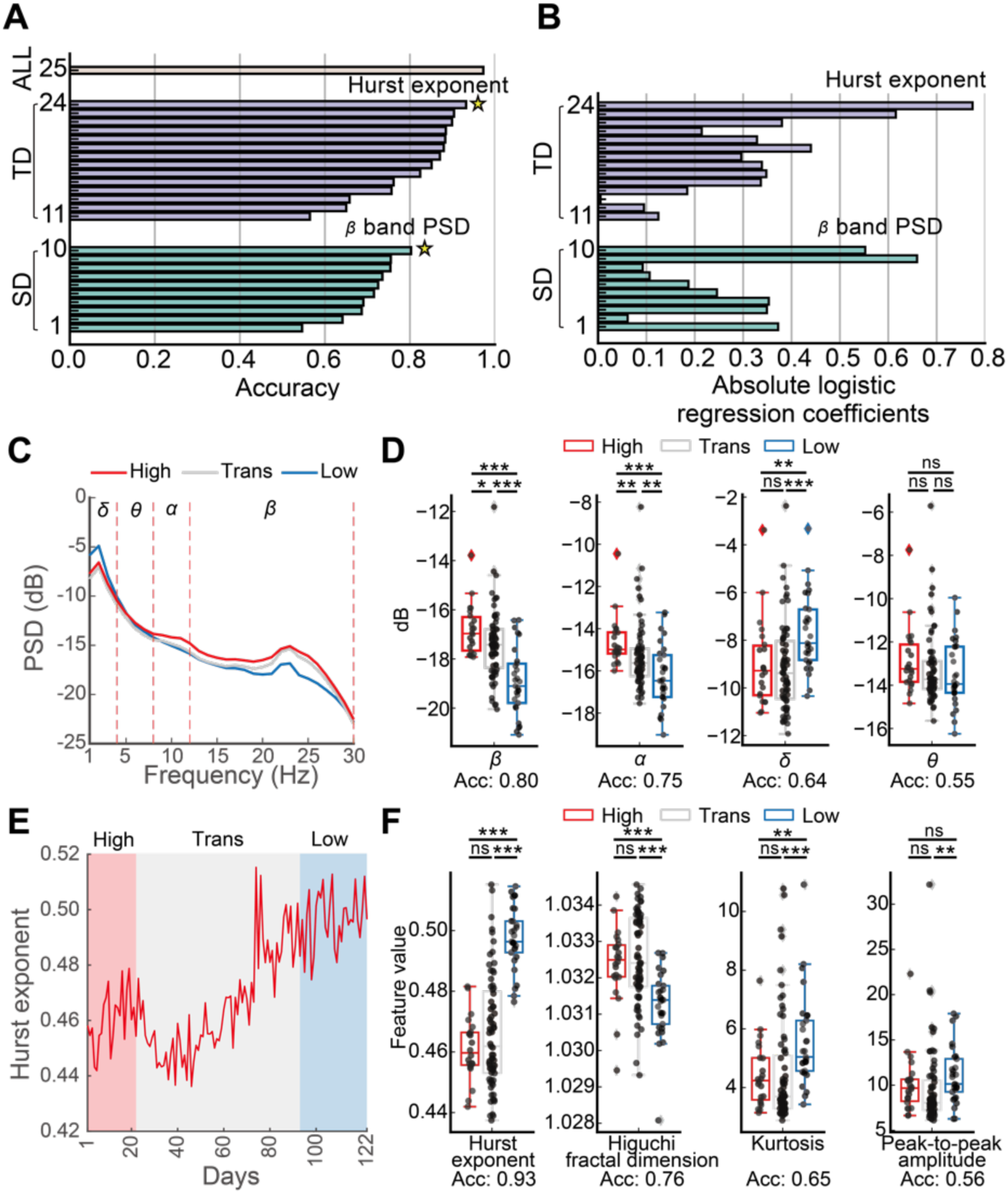
Importance of the *β* band spectral feature and the Hurst exponent temporal feature to the identification of neural biomarker and the tracking of the depression symptom state. (A) Classification accuracy of individual features and all LFP features by separately training LR models. The individual spectral domain (SD) features are in green, and their indices are ordered based on classification accuracy. The individual temporal domain (TD) features are in blue, and their indices are ordered by classification accuracy. The combination of all LFP features is in brown. The best individual feature was indicated by a yellow star for each domain. (B) The absolute coefficients of each feature in the LR model trained with all LFP features. The feature indices are the same as in (A). (C) Changes in the average LFP spectrum from the high symptom state (High) to the transition symptom state (Trans), and finally to the low symptom state (Low). (D) Changes in each of the four frequency bands with individual feature classification accuracy (Acc) indicated below. Two-sided Wilcoxon rank-sum test was used for significance test. Asterisks indicate the significance levels \**p*<0.05, \*\**p*<0.01, \*\*\**p*<0.001, and ns indicates no significance. (E) Changes in the temporal domain feature Hurst exponent from High to Trans to Low state. (F) Same as (D) but for four example temporal domain features.

We then examined the variations of *β* band PSD and Hurst exponent at the high, transition and low symptom states. In terms of the spectral domain, the overall LFP PSD showed obvious changes during the three symptom states (Figure 10C), with the *β* band PSD showing a significant and consistent decrease when changing from high state, to transition state, and finally to low state (Figure 10D). By contrast, no significant change was found for the *θ* band PSD and less consistent changes was found for the *δ* band and *α* band PSD (Figure 10D). In terms of the temporal domain, the Hurst exponent showed an increase when changing from high state, to transition state, and finally to low state (Figure 10E and 10F) while other representative temporal domain features showed less obvious changes (Figure 10F). Together, these results demonstrated the important contribution of the *β* band spectral feature and the Hurst exponent temporal feature to the identification of neural biomarker and the tracking of depression symptom state.

### 3.8 The identified neural biomarker and its contributing features indicated changes in LHb excitatory/inhibition (E/I) balance

We then interpreted the identified neural biomarker by relating it to the E/I balance of LHb activity. We used the LHb LFP spectrum’s 1/*f* slope as a possible indicator of LHb E/I balance, where a larger absolute 1/*f* slope indicates more inhibition (see Section 2.9). We found that after DBS treatment, the LHb activity changed to a more inhibitory state (Figure 11A). More specifically, the LHb E/I balance indicator increased when high symptom state changed to low symptom state (Figure 11B) and showed a trend of increase during the entire DBS treatment process (Figure 11C), consistently indicating an increase of inhibition. Moreover, we found a significant positive correlation between the identified neutral biomarker and the LHb E/I balance indicator (Figure 11D, Spearman’s *ρ* = 0.78, *P* = 2.7 × 10^-6^). Further, the most contributing Hurst exponent temporal feature and *β* band spectral feature both significantly correlated with the LHb E/I balance indicator (Figure 11D, Hurst exponent: Spearman’s *ρ* = −0.80, *P* = 1.9 × 10^-2’^; *β* band PSD: Spearman’s *ρ* = 0.20, *P* = 2.8 × 10^-2^). These results suggest that LHb E/I balance changed towards more inhibition following DBS treatment and our identified neural biomarker and its most contributing features significantly tracked such a change in LHb E/I balance.

**Figure 11.**
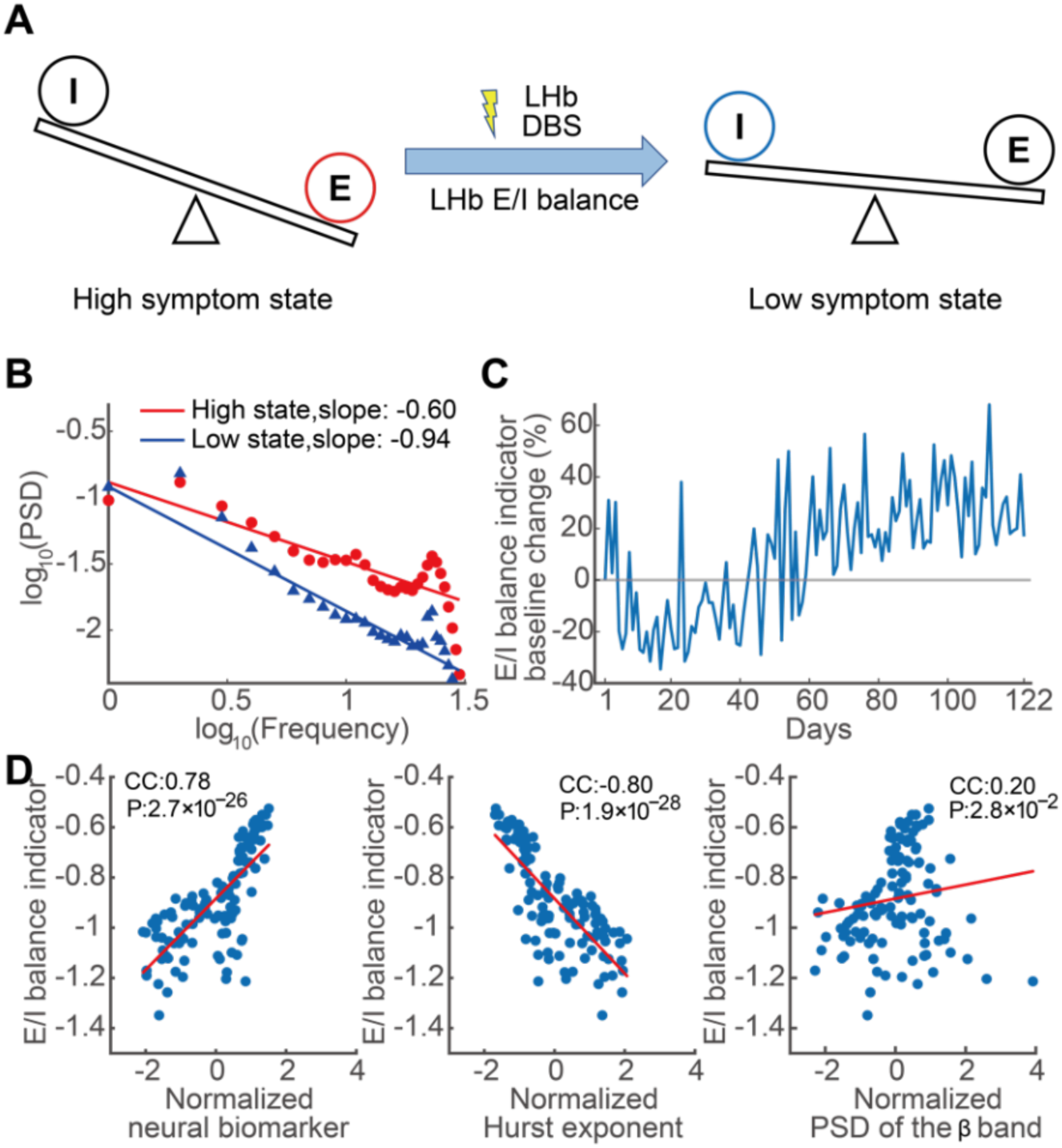
Changes of E/I balance during LHb DBS treatment and its’ correlations with the identified neural biomarker and the most contributing features. (A) Overview of the LHb E/I balance change after DBS treatment. (B) LHb LFP spectrum’s 1/*f* slope of two example days in high symptom state and low symptom state, respectively. The LFP spectrum’s 1/*f* slope is taken as the E/I balance indicator. (C) Temporal trace of baseline-normalized daily LHb E/I balance indicator. (D) Correlation between the E/I balance indicator and the identified neural biomarker (left), between the E/I balance indicator and the Hurst exponent (middle), and the E/I balance indicator and the *β* band PSD (right).

### 3.9 The DBS treatment involved a critical LHb-DRN circuit that possibly regulates depression via E/I balancing

After finding a connection between the neural biomarker and LHb E/I balance, we finally investigated the underlying neural circuits of LHb E/I balance. We used diffusion tensor imaging to obtain white matter fibers related to LHb and found that strong white matter fiber connections exist between LHb and the dorsal raphe nucleus (DRN) in our patient (Figure 12A).

**Figure 12.**
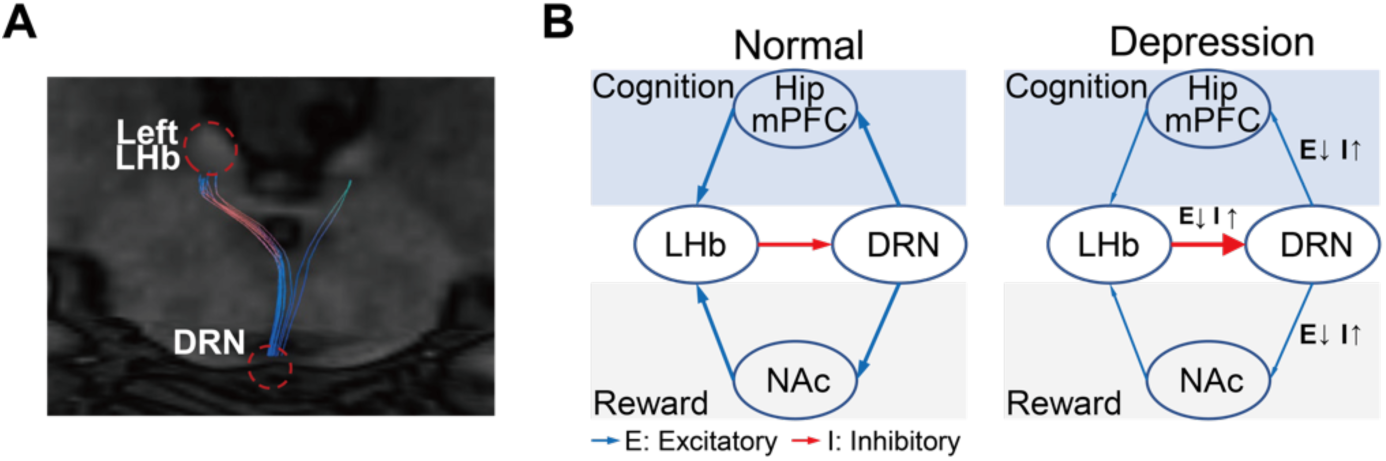
Diffusion tensor imaging of the white matter tracks related to LHb and a possible regulation pathway for depression. (A) Diffusion tensor imaging of the white matter tracks related to LHb. The DBS target (left LHb) and the connected brain area (DRN) were circled in red. Note that while the right LHb was not stimulated, its related white matter fibers are also shown. (B) LHb-DRN circuit and its possible role in regulating the cognition and reward dimensions of depression symptom. Hip: hippocampus; mPFC: medial prefrontal cortex; NAc: nucleus accumbens.

The LHb-DRN circuit is a critical pathway involved in central serotonergic regulation. This circuit plays a role in regulating cognition and reward, both of which are essential symptom dimensions associated with depression. The involvement of the LHb-DRN circuit suggests a possible mechanism of DBS treatment in our patient (also see Section 4.3). Specifically, prior studies have shown that in depressed patients, the LHb activity is overly excited (unbalanced E/I towards excitation), thus exerting substantial inhibition on the DRN via the inhibitory connections between LHb and DRN, consequently leading to reduced serotonin output from the DRN. Such a reduction in serotonin further inhibits the hippocampus and medial prefrontal cortex activity, possibly asserting influence on the cognitive dimension of depression; it also inhibits the nucleus accumbens activity, possibly asserting influence on the reward dimension of depression (Figure 12B). Our results suggest that LHb DBS treatment moved the LHb activity to a more inhibitory state, possibly restoring the LHb E/I balance towards a normal state and improving the depression symptoms (especially cognition functions and emotion blunting, see Section 3.1) via the critical regulation pathway involving LHb and DRN.

## 4. Discussion

### 4.1 Improvement of depression symptoms, emotional blunting, and cognitive function during DBS treatment

The clinical evaluation results show that our patient not only achieved improvements of HAMD, MADRS, and HAMA ratings that are typically used as clinical gold standard in prior DBS studies [11,13,17], but also achieved notable improvements of emotional blunting and cognition functioning. Almost three-quarters of patients in the acute phase of depression and one-quarter of those in remission reported severe emotional blunting. Approximately 56^7^ of patients considered their emotional blunting to be caused by their depression, while 45^7^ believed that their antidepressant medication was negatively affecting their emotions [63]. Emotional blunting has a substantial negative impact on patients’ daily functioning, well-being, and quality of life in both the acute and remission phases of depression [64]. On the other hand, cognitive deficits may be inherent to TRD and occur independently of affective symptomatology [65]. Objective cognitive assessment for the clinical evaluation for patients with TRD is important [66]. Thus, in our TRD patient, we specifically addressed the evaluation of both emotional blunting and cognitive functioning and showed improvements along both aspects.

However, our results regarding emotional blunting and cognitive functioning should be interpreted cautiously because of the lack of controlled data. Due to limited data about cognition in this case, we did not identify a neural biomarker to classify these cognitive states. The cognitive impact of DBS has usually been previously evaluated for epilepsy, movement disorder, and obsessive-compulsive disorder patients but rarely in controlled studies on TRD. Our study found no cognitive decline and suggests positive effects of DBS on cognitive functioning in TRD consistent with a previous report [67].

### 4.2 A data-driven LHb neural biomarker for tracking slow and fast depression symptom variations during DBS treatment

A mechanism-driven neural biomarker for depression is currently lacking mainly because the neural circuitry underlying depression has not been clearly delineated [68]. Therefore, current neural biomarkers of depression symptoms for tracking DBS effects have largely used data-driven machine learning methods to map LFP features to depression symptom ratings [17,32,33]. The usefulness of data-driven neural biomarkers critically depends on the type of data used to identify the neural biomarker. For example, a recent work [32] focused on a cingulate neural biomarker that was trained with and accordingly predicted longer-term (on the time scale of weeks) clinical ratings. On the other hand, another recent work [17] only trained and tested a neural biomarker with shorter-term (on the time scale of minutes) self-reports. Our work is unique in that, while we identified our LHb neural biomarker based on weekly clinical ratings, we demonstrated that the neural biomarker predicted not only weekly clinical ratings (data not used in identification) but also daily self-reports (data again not used in identification). The results suggest that our LHb neural biomarker could track the temporal dynamics of both slow and fast depression symptom variations, which was useful for developing new aDBS strategies that are robust across different time scales. It is worth noting that our LHb neural biomarker specifically tracked the temporal dynamics of weekly and daily depression symptom scores but not the anxiety symptom scores. It suggests that despite the overlapping of depression-related and anxiety-related brain networks [69], LHb neural activity is mainly related to depression, which is supported by prior animal studies [21–23].

Both population-level and personalized neural biomarkers of depression symptoms have been identified for tracking DBS effects. Population-level neural biomarkers are derived from data collected from several patients and have the benefits of being directly applicable to a new patient and robust interpretability of the neural biomarker’s biophysical mechanism across patients [32]. By contrast, personalized neural biomarkers are derived from data collected from an individual patient, which is more powerful in capturing the unique characteristics of depression symptoms in each patient, especially given the large inter-individual variability in depression-related brain networks [70]. With the emerging capability of recording more data within a single patient using mobile devices, personalized neural biomarker models can be more accurate in tracking the temporal dynamics of depression symptom variations. With such trends, a personalized neural biomarker has been identified and used for realizing aDBS targeting VC/VS [17]. Our study identified a personalized neural biomarker that achieved accurate classification and tracking of depression symptom variations during the DBS treatment targeting LHb, confirming the usefulness of personalization. Nevertheless, population-level and personalized neural biomarkers can complement each other. For example, one can leverage a large amount of population data to train an interpretable population-level neural biomarker model, followed by fine-tuning with personalized data to further improve its accuracy, which is an important future research direction.

A critical issue in identifying data-driven neural biomarker, especially personalized neural biomarker, is the collection of sufficient amounts of data. A recent population-level study collected LFP and clinical rating data from 5 individuals on a weekly basis over a span of 24 weeks [32], resulting in a total data sample size of 120, which was deemed sufficient for obtaining a neural biomarker at the population level. Several personalized studies collected LFP or iEEG and self-report data over several days, with a data sample size of around 30 for each individual [17,33]. By contrast, our study involved daily data collection over a much longer period of 41 weeks. Even after removing noisy data epochs, our data-driven neural biomarker analyses were based on a substantial data sample size of 122 days, which significantly exceeded the data sample size of one individual in the aforementioned studies. Our robustness analyses confirmed the sufficiency of our data sample size, showing that in our patient, the neural biomarker performance maintained stable even after reducing the data sample size to around 70 but had significant variations after reducing the data sample size below 40. This result indicates the importance of collecting sufficient data for identifying personalized neural biomarkers. The ongoing advancements in data recording devices offer exciting prospects for recording more data samples within an individual, thus facilitating the future development of personalized neural biomarkers.

### 4.3 Neurobiological interpretation of the identified neural biomarker and its contributing features in terms of LHb E/I balance

Previous studies have exclusively used LFP spectral domain features to identify neural biomarkers for depression [17,32,33]. By contrast, we found that both the LFP temporal domain and spectral domain features contributed to our neural biomarker and that the temporal domain features contributed relatively more than the spectral domain features. The main contributing temporal domain feature was the Hurst exponent. The Hurst exponent mainly indicates the stochasticity and predictability of a time-series [71]. We found that during the high and transition symptom states, the LHb LFP Hurst exponent values were mostly below 0.5, which indicated more stochastic and random-walk-like neural dynamics [72]. By contrast, during the low symptom state towards the end of DBS treatment, the LHb LFP Hurst exponent values were approaching and even surpassing 0.5, which indicated more predictable and stable neural dynamics [73]. Stochastic vs. predictable neural dynamics have been reported to relate with E/I balance of neural firing [74]. Related to this explanation, Hurst exponent has been widely used to examine E/I balance of the neural circuits in other neuropsychiatric disorders such as autism [75,76]. Our results suggest that a DBS-induced increase in LHb inhibition led to an increase in the Hurst exponent, moving LHb activity to a more E/I balanced state.

The main contributing spectral domain feature was the *β* band PSD. The LHb *β* band oscillation was also found to correlate with depression symptoms before DBS treatment in a previous study [28]. The *β* band oscillation might reflect the abnormal E/I balance of LHb neural ensembles underlying depression because it can depend on the firing patterns of a network of inhibitory interneurons gated by their mutually induced GABA_A_ action [77]. Moreover, LHb *β* band oscillation might be related to the abnormal burst spiking phenomena of LHb neurons found in rodents exhibiting depression-like behaviors [21–23]. These findings highlight the significance of the *β* band oscillations in LHb as related to depression.

Existing neuroscience findings provide evidence that LHb E/I balance influences the dopaminergic and serotoninergic projections from LHb and directly affects two neural pathways: the LHb-VTA (ventral tegmental area) pathway mainly regulates the dopaminergic activity, while the LHb-DRN pathway primarily regulates the serotoninergic activity [34]. Our white fiber tracking results predominantly reflected the influence of DBS on the LHb-DRN pathway. The DBS might have inhibited overly-excited LHb activity, activating overly-inhibited DRN activity, and restoring downstream neural activity related to cognition and reward circuits related to depression, which finally led to the multi-faceted improvement of depression symptoms, emotional blunting, and cognitive functioning of our patient. However, due to the complexity of LHb-related circuits, the exact neural activity changes of the LHb-related circuit in DBS treatment of depression requires further investigation by collecting multimodal neural data from more brain regions.

It is worth noting that due to the limitation of DBS electrode, we only used a simple LFP 1/*f* slope as an indirect indicator for LHb E/I balance. The E/I balance is directly related to the spiking activity of excitatory neurons and inhibitory neurons, and further related to the neuronal homeostasis and the formation of neural oscillations [78,79]. The underlying excitatory and inhibitory neuron synaptic currents mix together to give rise to the 1/*f*-like nature of the LFP PSD [34]. Previous study found that alterations of E/I balance within neural circuits can indeed be inferred from changes in the LFP 1/*f* slope [80]. Nevertheless, the LFP signal represents the collective activity of multiple neurons and only provides an indirect measure of E/I balance. Future work can use more advanced micro-macro electrodes [81] to record LHb spiking activity to more directly quantify LHb E/I balance.

### 4.4 DBS frequency deferentially modulates the depression symptom and neural biomarker

Prior studies have shown that DBS frequency can significantly influence the treatment efficacy for TRD, e.g., high-frequency DBS has generally yielded better treatment outcomes than low-frequency DBS [24,82,83]. Consistently, we discovered that a very low DBS frequency of 1 Hz was not effective in alleviating the depression symptoms in our patient, but higher frequencies of 20 Hz and 130 Hz were more effective. Beyond the depression symptom ratings, we additionally found that the neural biomarker was also consistently modulated by the different DBS frequencies. DBS frequency might influence the release of neurotransmitters in depression-targeted pathways [84], thus modulating the neural biomarker and the depression symptoms. However, similar to other DBS targets, the optimal DBS frequency at LHb is still also an open question that requires further research.

## 5. Limitations

Our study has several limitations. First, while our study had a sufficient within-patient data sample size, the patient sample size was limited (n-of-1); further studies with more patients are needed to confirm our findings on LHb neural biomarkers. Second, despite its powerful classification and tracking performance, our neural biomarker was identified using only one channel of LFP signals due to the limitation of device hardware configuration. Incorporating multi-channel LFP signals in future studies would allow for finding neural biomarkers with even better performance and a more comprehensive understanding of the neural mechanism underlying neural biomarker identification. Third, due to the high-frequency recording noise of our DBS device, we filtered the LFP signal below 30 Hz to ensure noise rejection. Future work with better recording capability should investigate how higher-frequency LFP temporal and spectral domain features contribute to the identification of neural biomarkers. Finally, to remove the confounding factor of stimulation artifact, when we recorded LFP, we completely turned off stimulation. While this approach ensured stimulation-artifact-free LFP signals, it is important to consider using LFP signals during stimulation to identify better responsive neural biomarkers in the future, but this requires high-performance stimulation artifact removal, which remains challenging [85].

## 6. Conclusion

One patient with TRD reached remission after 41 weeks of LHb DBS treatment. With a unique longitudinal data collection of concurrent daily and weekly depression symptom scores and LHb LFP signals during the entire treatment process, we used machine learning to identify an LHb neural biomarker of depression symptoms, with the most contributing spectral feature being LFP *β* band power and temporal feature being Hurst exponent. We demonstrated that our LHb neural biomarker accurately classified high and low depression symptom severity states, simultaneously tracked the temporal dynamics of weekly (slow) and daily (fast) depression symptom variations during the DBS treatment process, and reflected the depression symptom changes in response to DBS frequency alterations. We also interpreted the neural biomarker as indicating changes in LHb excitatory/inhibition balance during DBS treatment. Our methods and results hold promise in identifying clinically-viable neural biomarkers to facilitate future adaptive DBS developments for treating TRD.

## Data availability

The data supporting this study’s findings are available from the corresponding author upon reasonable request.

## Author Contributions

Shi Liu: Methodology, Software, Formal analysis, Writing - Original Draft, Visualization. Yu Qi: Investigation, Resources. Shaohua Hu: Conceptualization, Resources. Ning Wei: Resources, Data Curation, Writing - Original Draft. Jianmin Zhang: Conceptualization, Resources. Junming Zhu: Conceptualization, Resources. Hemmings Wu: Conceptualization, Investigation. Hailan Hu: Conceptualization. Yuxiao Yang: Conceptualization, Supervision, Methodology, Writing - Review ^6^ Editing, Funding acquisition. Yueming Wang: Conceptualization, Supervision, Project administration, Funding acquisition, Writing - Review ^6^ Editing. All authors revised and approved the final version of the manuscript.

