## Supplemental files for "A Habenula Neural Biomarker Simultaneously Tracks Weekly and Daily Symptom Variations during Deep Brain Stimulation Therapy for Depression"

**Table S1. The summary of the extracted 24 features from LHb LFP signals. The spectral and temporal domain features are ordered separately based on classification accuracy and finally summarized together.**

| **ID** | **Feature Name (Abbr.)** | **Domain** | **Explanation** |
| --- | --- | --- | --- |
| **1** | **psd_theta** | **SD** | PSD of theta band |
| **2** | **psd_delta** | **SD** | PSD of delta band |
| **3** | **pac_4_8c12_30** | **SD** | PAC, phase band: 4-8Hz,  amplitude band: 12-30Hz |
| **4** | **pac_8_12c12_30** | **SD** | PAC, phase band: 8-12Hz,  amplitude band: 12-30Hz |
| **5** | **pac_1_4c12_30** | **SD** | PAC, phase band: 1-4Hz,  amplitude band: 12-30Hz |
| **6** | **pac_4_8c8_12** | **SD** | PAC, phase band: 4-8Hz,  amplitude band: 8-12Hz |
| **7** | **pac_1_4c4_8** | **SD** | PAC, phase band: 1-4Hz,  amplitude band: 4-8Hz |
| **8** | **pac_1_4c8_12** | **SD** | PAC, phase band: 1-4Hz,  amplitude band: 8-12Hz |
| **9** | **psd_alpha** | **SD** | PSD of alpha band |
| **10** | **psd_beta** | **SD** | PSD of beta band |
| **11** | **ptp_amp** | **TD** | peak-to-peak amplitude |
| **12** | **kurtosis** | **TD** | kurtosis |
| **13** | **skewness** | **TD** | skewness |
| **14** | **katz_fd** | **TD** | katz fractal dimension |
| **15** | **higuchi_fd** | **TD** | Higuchi fractal dimension |
| **16** | **samp_entropy** | **TD** | sample entropy |
| **17** | **app_entropy** | **TD** | approximate entropy |
| **18** | **svd_entropy** | **TD** | singular value decomposition entropy |
| **19** | **zero_crossings** | **TD** | zero-crossings |
| **20** | **svd_fisher_info** | **TD** | singular value decomposition fisher information |
| **21** | **hjorth_mobility** | **TD** | Hjorth mobility |
| **22** | **hjorth_complexity** | **TD** | Hjorth complexity |
| **23** | **line_length** | **TD** | line length |
| **24** | **hurst_exp** | **TD** | Hurst exponent |

***SD*** Frequency domain, ***TD*** Time domain.

**Table S2. Spearman’s rank correlation coefficients between weekly clinical ratings (HAMD, HAMA, MADRS) and daily self-reports (VAS-D, VAS-A).**

| **Scales** | **HAMD** | **HAMA** | **MADRS** |
| --- | --- | --- | --- |
| **VAS-D** | **0.931** | **0.730** | **0.880** |
| **VAS-A** | **0.703** | **0.695** | **0.734** |

**Table S3. Comparisons of the performance of six different classifiers.**

| **Classifier** | **Accuracy** | **Specificity** | **Sensitivity** | **F1-score** | **AUC** |
| --- | --- | --- | --- | --- | --- |
| **LR** | **0.973** | **0.961** | **0.988** | **0.970** | **0.974** |
| **MLP** | 0.950 | 0.947 | 0.953 | 0.942 | 0.950 |
| **AdaBoost** | 0.950 | 0.950 | 0.952 | 0.942 | 0.951 |
| **SVM** | 0.947 | 0.943 | 0.954 | 0.940 | 0.948 |
| **RF** | 0.937 | 0.959 | 0.909 | 0.924 | 0.934 |
| **LDA** | 0.912 | 0.878 | 0.956 | 0.906 | 0.917 |

LR = Logistic regression; MLP = Multilayer perceptron; AdaBoost = Adaptive Boosting; SVM = Support vector machine; RF = Random Forest; LDA = Linear discriminant analysis.

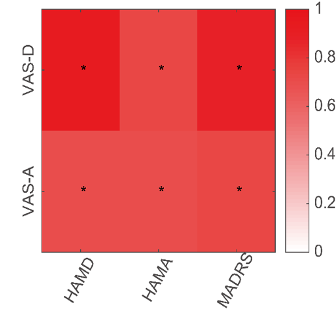

**Figure S1.** Heatmap of the correlation coefficients between weekly clinical ratings (HAMD, HAMA, MADRS) and daily self-reports (VAS-D, VAS-A). Cells marked with * indicate the coefficients that are significantly different from zero (Bonferroni corrected *p* < 0.05).

**Note S1. Patient medical information**

This study included a male TRD patient aged 36-40 years old. The patient suffered from depression beginning before the age of 18 without any hypomanic or manic episodes. In the past 19 years, the patient has experienced severe depressive symptoms accompanied by cognitive deficits, anhedonia, and impaired social functioning. Despite undergoing multiple pharmacotherapeutic treatments involving various antidepressants and augmentation strategies with selective serotonin reuptake inhibitors (SSRIs), serotonin and norepinephrine reuptake inhibitors (SNRIs), as well as other antipsychotic medications, the patient's depression symptoms remained uncontrolled, leading to significant functional impairment in recent years.

**Note S2. Stimulation parameters during DBS treatment process**

We divided the treatment process into six stages based on the alterations of stimulation parameters: 1) The “Preop” stage, starting in November 2021 and lasting 37 days, was the time before the DBS electrode implantation. 2) The “Off-1” stage lasted 22 days, during which the patient recovered from surgery and stimulation was not activated. 3) The “1 Hz” stage lasted 35 days, during which we set the stimulation frequency at 1 Hz using a pulse width of 90 μs and a voltage of 4 V. 4) The “Off-2” stage lasted 34 days, during which the DBS device shut down because of an unnoticed power off. 5) The “20 Hz” stage lasted 103 days, during which we re-started DBS and set the stimulation frequency at 20 Hz using a pulse width of 90 μs and a voltage of 4 V. 6) The “130 Hz” stage lasted 93 days, during which we set the stimulation frequency to 130 Hz using a pulse width of 90 μs and a voltage of 3.5 V.

**Note S3. Removal of LFP signal bad epochs and feature extraction**

To remove bad epochs from daily LFP signals, we computed the power spectral density (PSD) of four frequency bands: $\delta$(1-4 Hz), $\theta$(4-8 Hz), $\alpha$(8-12 Hz), and $\beta$(12-30 Hz) to identify noisy epochs with outlier PSD features in any band (removed using the Matlab function rmoutliers with the quartiles method.) The remaining epochs were used for further feature extraction.

For each remaining LFP epoch, we computed its spectral domain (SD) and temporal domain (TD) features. For the SD features, we calculated the PSD of the four frequency bands mentioned above, quantifying the oscillations within the specific frequency bands. We used the *pwelch* function in Matlab with a Hamming window with a size of 1000 and an overlap size of 500. We also calculated the phase-amplitude coupling (PAC), which quantifies the coupling relationship between low-frequency phase and high-frequency amplitude. We calculated the PAC with the standard cross-frequency coupling algorithm used in previous studies [6,7]. The bandwidth was set to the width of the frequency range of interest. Six pairs of coupling were estimated: phase band vs. amplitude band: 1-4 Hz vs. 4-8 Hz, 1-4 Hz vs. 8-12 Hz, 1-4 Hz vs. 12-30 Hz, 4-8 Hz vs. 8-12 Hz, 4-8 Hz vs. 12-30 Hz, 8-12 Hz vs. 12-30 Hz. (2) For TD features, we used the *mne_features* python library to calculate fourteen temporal domain features using the default parameters [8], including the approximate entropy, Higuchi fractal dimension, Hjorth complexity, Hjorth mobility, Hurst exponent, Katz fractal dimension, Kurtosis, line length, peak-to-peak amplitude, sample entropy, skewness, singular value decomposition entropy, singular value decomposition Fisher information and zero-crossings.
